# Antivirals for treatment of severe influenza: a systematic review and network meta-analysis of randomized controlled trials

**DOI:** 10.1101/2024.05.28.24307938

**Authors:** Ya Gao, Gordon Guyatt, Timothy M Uyeki, Ming Liu, Yamin Chen, Yunli Zhao, Yanjiao Shen, Jianguo Xu, Qingyong Zheng, Zhifan Li, Wanyu Zhao, Shuyue Luo, Xiaoyan Chen, Jinhui Tian, Qiukui Hao

## Abstract

**Background:** The optimal antiviral drug for treatment of severe influenza remains unclear. To support updated WHO influenza clinical guidelines, this systematic review and network meta-analysis evaluated antivirals for treatment of patients with severe influenza.

**Methods:** We systematically searched Medline, Embase, CENTRAL, CINAHL, Global Health, Epistemonikos, and ClinicalTrials.gov for randomized controlled trials published through 20 September 2023, that enrolled hospitalized patients with suspected or laboratory-confirmed influenza and compared direct-acting influenza antivirals against placebo, standard care, or another antiviral. We conducted frequentist network meta-analyses to summarize the evidence and evaluated the certainty of evidence using the GRADE (Grading of Recommendations Assessment, Development and Evaluation) approach. We registered the protocol with PROSPERO, CRD42023456650.

**Findings:** Of 11,878 records, 8 trials with 1,424 participants were included. The effects of oseltamivir, peramivir or zanamivir on mortality compared with placebo or standard care without placebo for seasonal and zoonotic influenza are uncertain. Compared with placebo or standard care, oseltamivir (mean difference (MD) 1.63 days lower, 95% CI 2.81 lower to 0.45 lower) and peramivir (MD 1.73 days lower, 95% CI 3.33 lower to 0.13 lower) may reduce duration of hospitalization for seasonal influenza (low certainty evidence). There were few or no differences between oseltamivir (MD 0.34 days higher, 95% CI 0.86 lower to 1.54 higher; low certainty evidence), peramivir (MD 0.05 days lower, 95% CI 0.69 lower to 0.59 higher; low certainty evidence) and standard care in time to alleviation of symptoms. There were no differences in adverse events or serious adverse events among oseltamivir, peramivir and zanamivir (very low certainty evidence).

**Interpretation:** In hospitalized patients with severe influenza, oseltamivir and peramivir may reduce duration of hospitalization compared with standard care or placebo. The effects of all antivirals on mortality and other important patient outcomes are very uncertain.

**Funding:** WHO.

**Research in context:** *Evidence before this study:* Antivirals are frequently used in the clinical management of people with severe influenza. Previous systematic reviews and meta-analyses have reported that early initiation of neuraminidase inhibitor (NAI) treatment in hospitalized influenza patients may be associated with reduced mortality and length of hospital stay compared with later or no NAI treatment. However, these pairwise meta-analyses mainly focused on the relative effects of one specific class of antivirals (NAIs), did not evaluate the effects of antivirals on severe zoonotic influenza, and did not assess the certainty of evidence. No network meta-analysis has evaluated all available antiviral treatments for severe influenza. The optimal antiviral drug for treatment of patients with severe influenza remains uncertain.

*Added value of this study:* We found low certainty evidence that oseltamivir and peramivir may reduce the duration of hospitalization in patients with severe seasonal influenza compared with placebo or standard care. Great uncertainty remains regarding the effects of oseltamivir, peramivir, and zanamivir on mortality in patients with severe seasonal influenza or zoonotic influenza. There are no important differences in adverse events or serious adverse events associated with oseltamivir, peramivir, or zanamivir for treatment of patients with severe influenza, although the evidence is of very low certainty. The effects of other antivirals, including baloxavir, in patients with severe influenza, on mortality and other important patient outcomes are very uncertain.

*Implications of all the available evidence:* Our study provides evidence that oseltamivir and peramivir, relative to placebo or standard care, may reduce the duration of hospitalization for patients with severe seasonal influenza. These findings primarily highlight the uncertainty regarding effects of antivirals for treatment of patients with severe influenza but do provide some justification for their use.

## Introduction

Influenza, a viral respiratory disease, typically causes mild to moderate upper respiratory symptoms that resolve within a week.^1–3^ However, a substantial proportion of individuals, particularly those in high-risk groups such as young children, older adults, pregnant women, and persons with chronic medical conditions, can develop severe illness from influenza.^1,4^

Influenza is an important cause of respiratory viral disease among hospitalized patients, with an estimated hundreds of thousands of respiratory deaths worldwide annually with major economic losses.^5–8^ Hospitalized patients with seasonal influenza may develop complications, including severe pneumonia, respiratory failure, multi-organ failure, and secondary bacterial infections, that can lead to death.^1,9–12^ The case fatality proportion for adults hospitalized with influenza typically ranges from 4% to 8%, but may be higher (>10-15%) during rare pandemics and among immunocompromised individuals.^13^ Therefore, identifying effective therapies for severe influenza is of global public health importance.

Antivirals, such as neuraminidase inhibitors (NAIs), are recommended and administered to patients with severe influenza.^14^ Systematic reviews and meta-analyses have reported that early NAI treatment may be associated with reduced mortality and shorter length of hospital stay compared with later or no NAI treatment for hospitalized influenza.^15–19^ However, these pairwise meta-analyses focused primarily on the relative effects of one class of antivirals (NAIs) for treatment of severe seasonal or pandemic influenza and did not assess effects of antivirals on zoonotic influenza nor assess the certainty of evidence.^15–19^ No network meta-analysis has evaluated all available antiviral treatments for severe influenza. The optimal antiviral drug for treatment of hospitalized patients with influenza remains uncertain.

To support an update of the World Health Organization (WHO) clinical guidelines for influenza,^20^ we performed a systematic review and network meta-analysis of randomized controlled trials (RCTs) to assess the efficacy and safety of antivirals for severe influenza.

## Methods

We registered this systematic review protocol with PROSPERO (CRD42023456650) and reported the review according to the guideline of Preferred Reported Items for Systematic Reviews and Meta-Analyses (PRISMA) for network meta-analyses.^21^

### Study selection and selection criteria

With the aid of a medical librarian, we searched Medline, Embase, Cochrane Central Register of Controlled Trials (CENTRAL), Cumulative Index to Nursing and Allied Health Literature (CINAHL), Global Health, Epistemonikos, ClinicalTrials.gov from databases inception through 20 September 2023 (Appendix 1) and reviewed reference lists of relevant systematic reviews to identify additional trials.

Eligible RCTs enrolled hospitalized patients with suspected or laboratory-confirmed influenza (confirmed by RT-PCR assay, rapid antigen test, or immunofluorescence assay) and compared direct-acting antivirals against placebo, standard care without placebo, or another antiviral for treatment of severe influenza. Severe influenza was defined by the WHO as an illness with laboratory-confirmed influenza that requires hospitalization.^20^ We focused on antivirals approved for treatment of influenza by the US Food and Drug Administration (FDA) or other regulatory organizations worldwide, including baloxavir, oseltamivir, laninamivir, zanamivir, peramivir, umifenovir, favipiravir, amantadine, and rimantadine.^22^ We did not apply restrictions on the type or subtype of influenza virus, publication language, patient age, or dose and administration route of antivirals but excluded studies that investigated influenza prevention with vaccines, Chinese medicines, antivirals combined with adjunctive therapies, or antivirals used for pre-or post-exposure chemoprophylaxis.

Using Covidence (https://covidence.org/), pairs of reviewers independently screened titles and abstracts of all citations and full texts of potentially eligible records. We checked retractions for all eligible publications; if a study was retracted, we excluded the study from our review.^23^ Pairs of reviewers independently extracted data on study characteristics, patient characteristics, characteristics of antivirals, and outcomes (Appendix 2.1). Reviewers resolved discrepancies by discussion or, if necessary, with the assistance of a third party for adjudication.

### Data analysis

The WHO guideline panel identified important patient outcomes as follows: time to alleviation of symptoms, duration of hospitalization, admission to intensive care unit (ICU), progression to invasive mechanical ventilation, duration of mechanical ventilation, mortality, hospital discharge destination, emergence of antiviral resistance, adverse events, adverse events related to treatments, and serious adverse events. We defined time to alleviation of symptoms as the duration between the start of treatment and the point at which influenza-associated symptoms are alleviated.^24,25^

Using the Hartung-Knapp-Sidik-Jonkman (HKSJ) random-effects model, we conducted pairwise meta-analyses for each direct comparison. For dichotomous outcomes, we calculated risk ratios (RRs) with 95% confidence intervals (CIs) for mortality, progression to invasive mechanical ventilation, emergence of resistance, any adverse events, adverse events related to treatments, and serious adverse events and risk differences (RDs) with 95% CIs for ICU admission. We calculated mean differences (MDs) with 95% CIs for continuous outcomes. When standard deviations (SDs) were missing, we estimated them using the methods described in the Cochrane Handbook.^26^ To assess the between-study heterogeneity, we used the I^2^ statistic and visually inspected forest plots. For comparisons that included at least 10 studies, to assess publication bias, we planned to use Harbord’s test for dichotomous outcomes and Egger’s test for continuous outcomes,^27,28^ as well as a visual assessment of the funnel plot.

We drew network plots for outcomes using STATA 15.0 (StataCorp, College Station, Texas, USA). We conducted frequentist random-effects network meta-analyses employing a graph-theoretical approach, with the estimator derived from weighted least-square regression using the Moore-Penrose pseudoinverse method. ^29^ Employing the “design-by-treatment” model (global test), we assessed the coherence assumption for the entire network.^30^ We calculated indirect estimates from the network by node-splitting and a back-calculation method.^31^ To assess local (loop-specific) incoherence within each closed loop of the network, measuring the difference between direct and indirect evidence, we applied the node-splitting method and computed a p-value for the incoherence test.^32^ We conducted the analyses in R version 4.2.1 (R Foundation for Statistical Computing).

To facilitate interpretation of results, we calculated absolute effects using RR estimates and the baseline risk estimates for outcomes in which the summary measure was RR. To estimate absolute effects of antivirals on mortality, the WHO guideline panel recommended use of two baseline risk categories for severe seasonal influenza and zoonotic influenza. We obtained baseline risks of mortality for severe seasonal influenza (30 per 1000) and zoonotic influenza [e.g., avian influenza A(H5N1), A(H5N6), or A(H7N9) virus infections] (387 per 1000) from meta-analyses (results will be reported elsewhere). For other outcomes for which reliable observational data were not available, we used the median baseline risk in the control group of eligible RCTs.

If data were available (at least two trials providing relevant information for each subgroup), we planned to perform the following prespecified within-trial subgroup analyses for patients with severe influenza:

1. Severe influenza etiology: seasonal influenza A and B viruses versus zoonotic influenza A viruses versus pandemic influenza A viruses (hypothesis: antiviral treatment has lower effectiveness in patients with zoonotic influenza than for seasonal or pandemic influenza).
2. Confirmed versus suspected influenza virus infection: (hypothesis: reduced treatment effect in patients with suspected influenza compared to patients with laboratory-confirmed influenza).
3. Age: children aged <2 years, children versus adults and adolescents versus elderly (aged ≥65 years) (hypothesis: reduced treatment effect in elderly).

We planned to assess the credibility of significant subgroup effects using the Instrument for assessing the Credibility of Effect Modification Analyses (ICEMAN) tool.^33^

Pairs of reviewers independently evaluated the risk of bias of eligible RCTs using a modified Cochrane risk of bias tool (Appendix 2.2).^34^

We used the Grading of Recommendations Assessment, Development, and Evaluation (GRADE) approach to assess certainty of evidence.^35,36^ By considering the risk of bias, inconsistency, indirectness, imprecision, publication bias, intransitivity, and incoherence, we rated certainty of evidence for each comparison and outcome as high, moderate, low, or very low.^37,38^ To assess intransitivity, we examined the distribution of potential effect modifiers, including age, influenza virus etiology, and confirmed or suspected influenza, across treatment comparisons. We assessed imprecision at the network level using the minimally important difference (MID) for an outcome as a threshold.^39^ The WHO guideline panel specified an MID of 0.3% for mortality, 1.5% for progression to invasive mechanical ventilation, 1% for admission to ICU, 1% for any adverse events and adverse events related to treatments, 0.5% for serious adverse events, 5% for emergence of antiviral resistance, and 1 day each for duration of hospitalization, time to alleviation of symptoms, and duration of mechanical ventilation. We rated imprecision following GRADE guidance.^40^ If incoherence was present, we used the estimate with the higher certainty of direct and indirect evidence as the best estimate. We developed the summary of findings tables in MAGICapp following GRADE guidance.^41,42^

## Results

Our search identified 11878 citations. After screening 8944 titles and abstracts and 459 full texts, 8 RCTs^43–50^ proved eligible (Figure 1).

**Figure 1.**
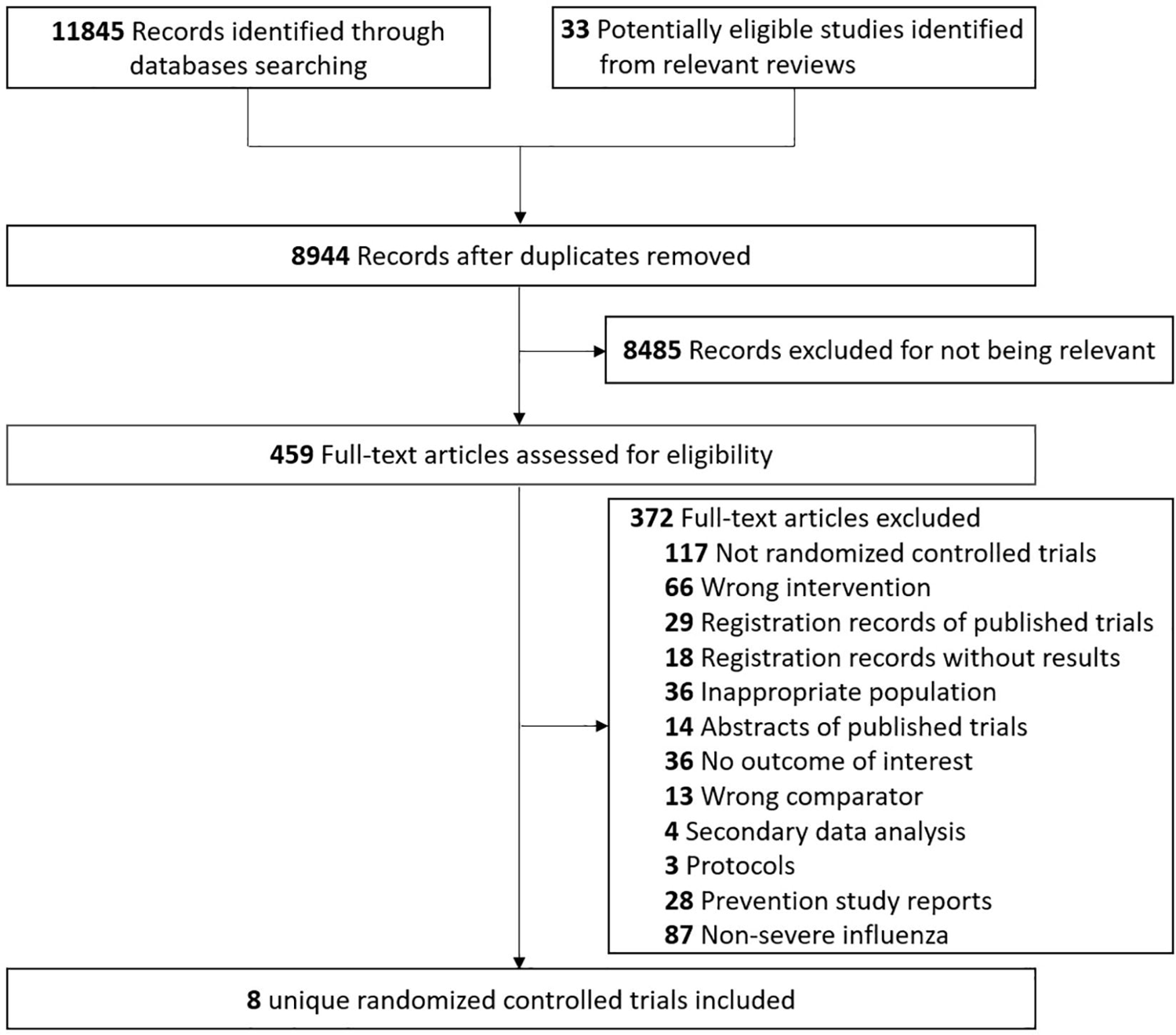
Study selection

### Study characteristics

The 8 eligible trials included a total of 1424 participants (ranging from 30 to 615 per trial). The mean age ranged from 20.5 to 60.3 years, the proportion of men ranged from 42.5% to 78.1%, and the proportion of laboratory-confirmed influenza patients ranged from 78% to 100%. The interventions included oseltamivir, peramivir, zanamivir, rimantadine, zanamivir plus rimantadine, and baloxavir plus NAIs. Direct comparisons between antivirals and standard care or placebo were available for oseltamivir and peramivir in three trials. The other five trials compared different antivirals: two trials compared oseltamivir against peramivir, one compared oseltamivir against zanamivir, one compared zanamivir and rimantadine against rimantadine alone, while the other trial compared baloxavir plus various NAIs against NAIs alone (Table 1 and Appendix 3).

**Table 1.**
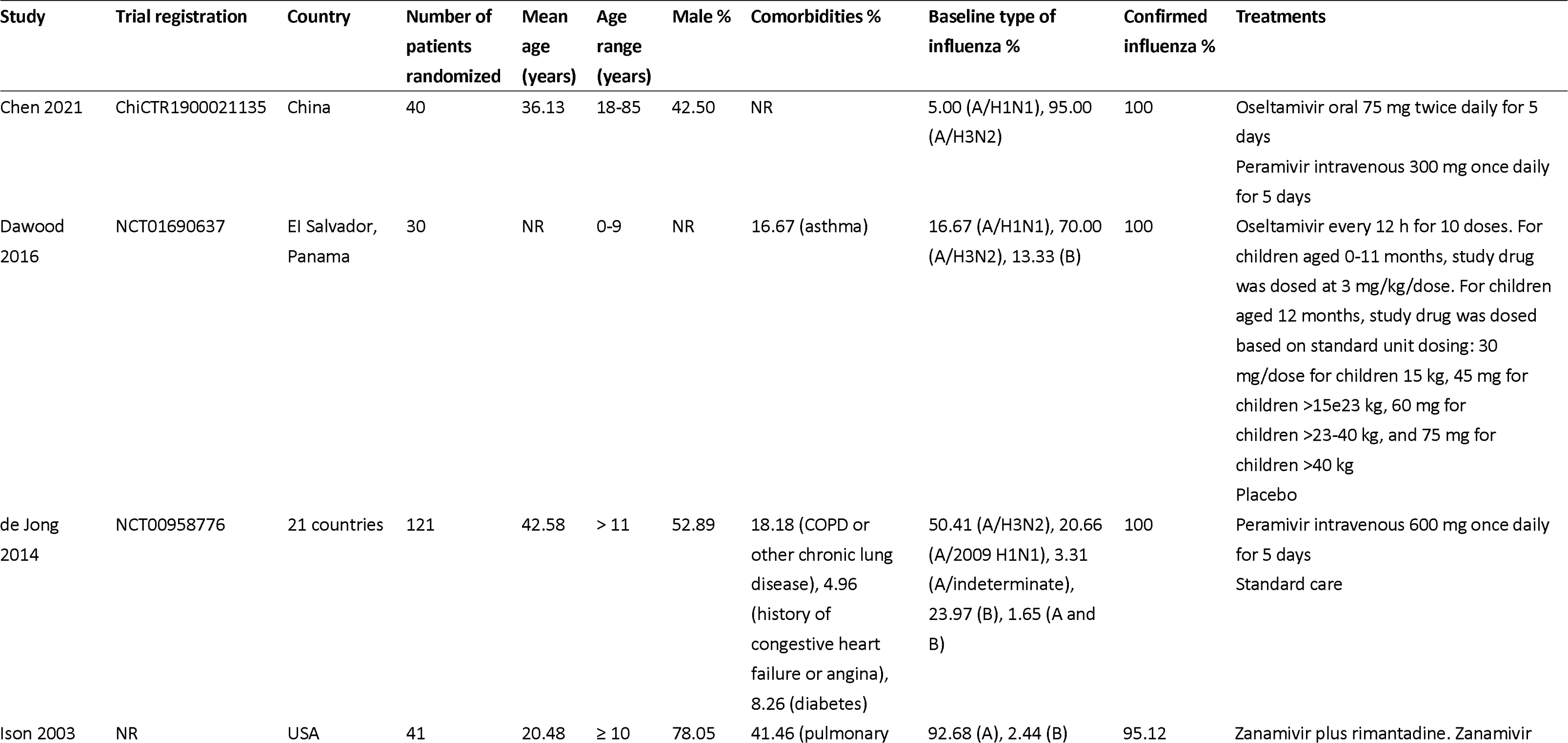
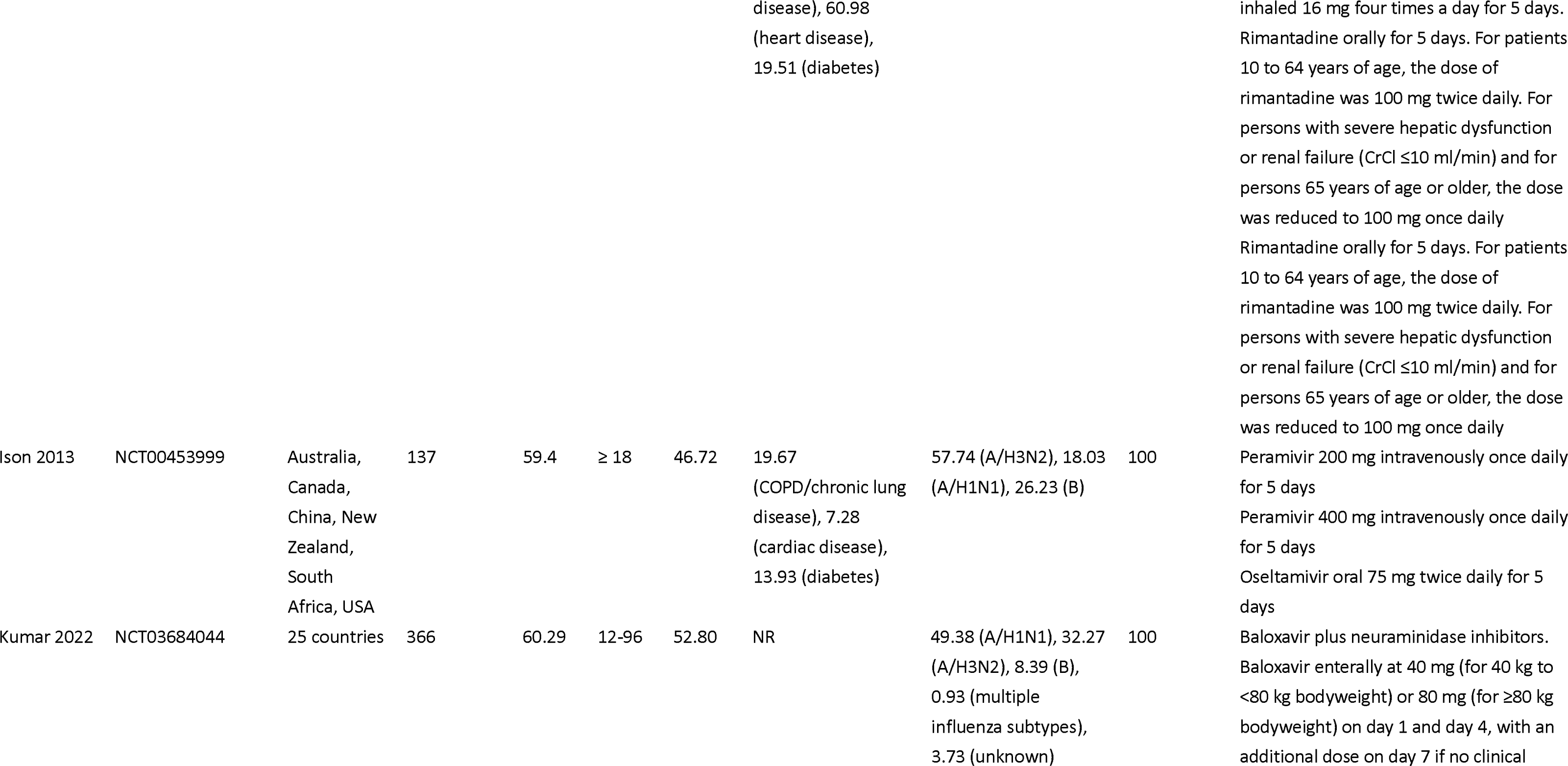
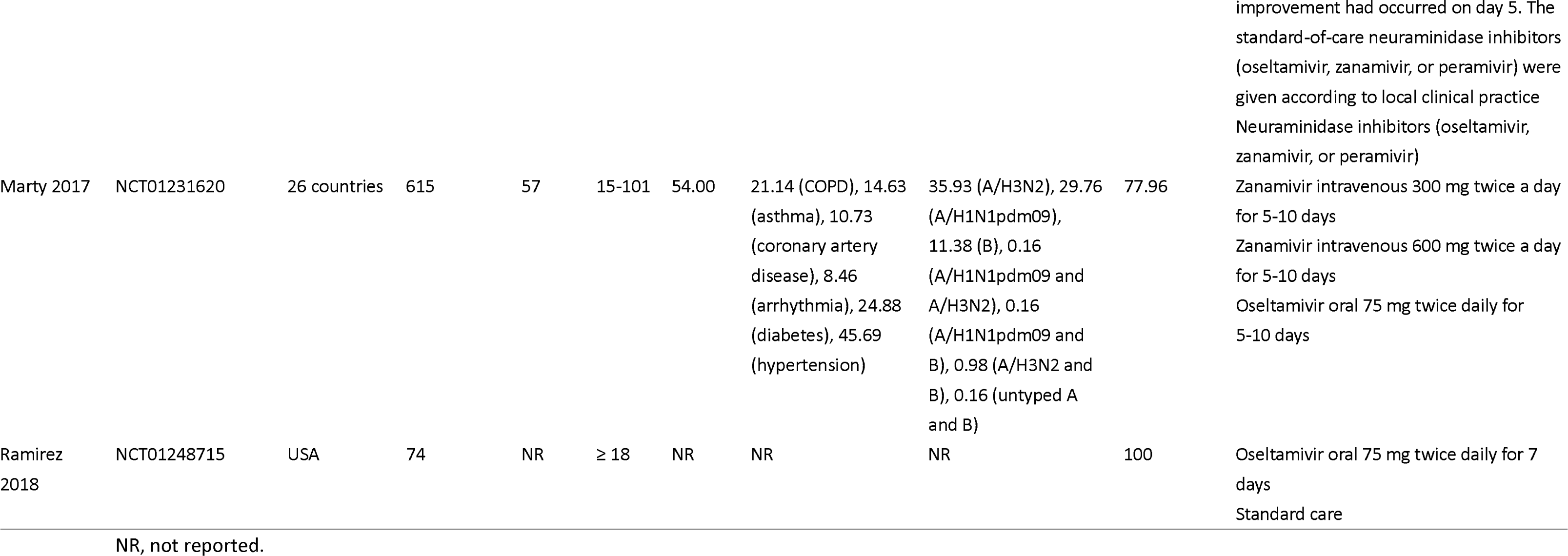
Basic characteristics of eligible RCTs.

### Risk of bias

Appendix 4 presents the risk of bias of eligible trials for each outcome. Most biases were due to inadequate allocation concealment and lack of blinding. We rated one trial as low or probably low risk of bias for all reported outcomes.^48^

### Network meta-analysis

Six trials were included in the network meta-analysis.^43–45,47,49,50^ Figure 2 and Appendix 5 present network plots for each outcome. We did not find substantial between-study heterogeneity (Appendix 6), global incoherence (Appendix 7), or local incoherence (Appendix 8). Table 2, Table 3, and Appendix 9 present GRADE summary of findings. We judged the certainty of evidence to be low or very low for all outcomes. We did not include two eligible trials in the network meta-analysis because both arms of these two trials did not connect with other interventions in the network.^46,48^

**Figure 2.**
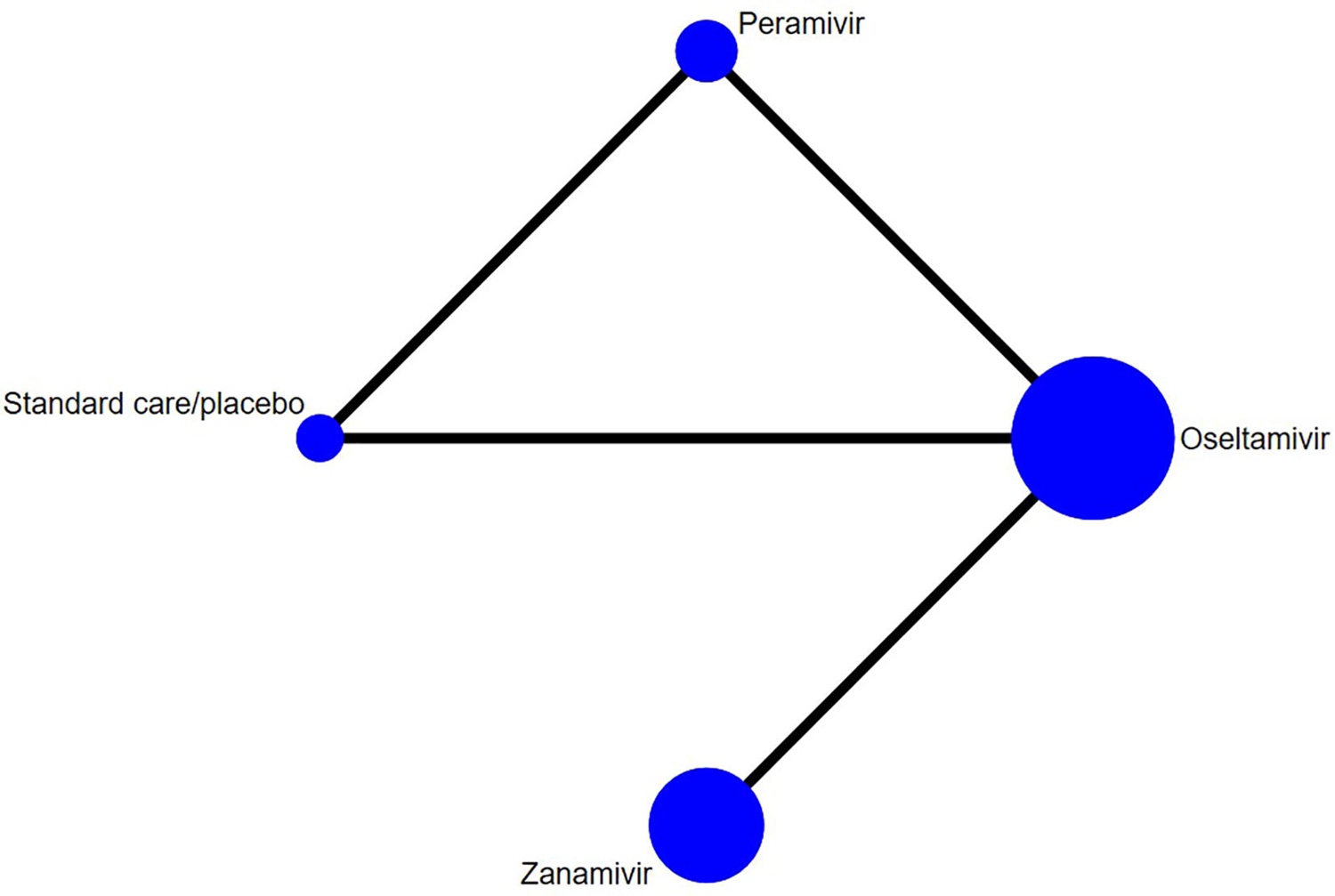
Network plot for mortality Footnotes: The size of the circle represents the number of participants. The connecting lines represent direct comparisons. The width of the line represents the number of studies.

**Table 2.**
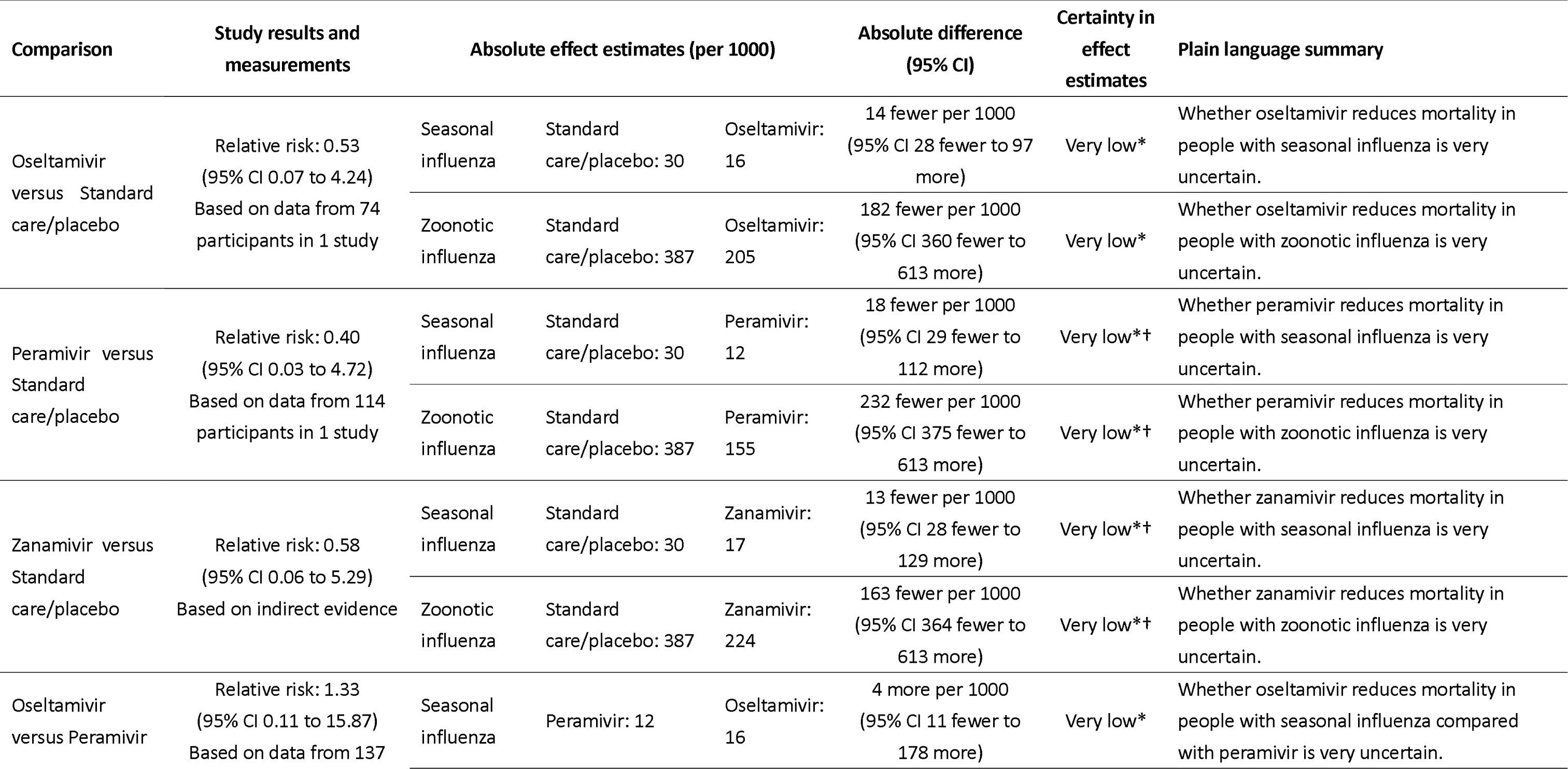
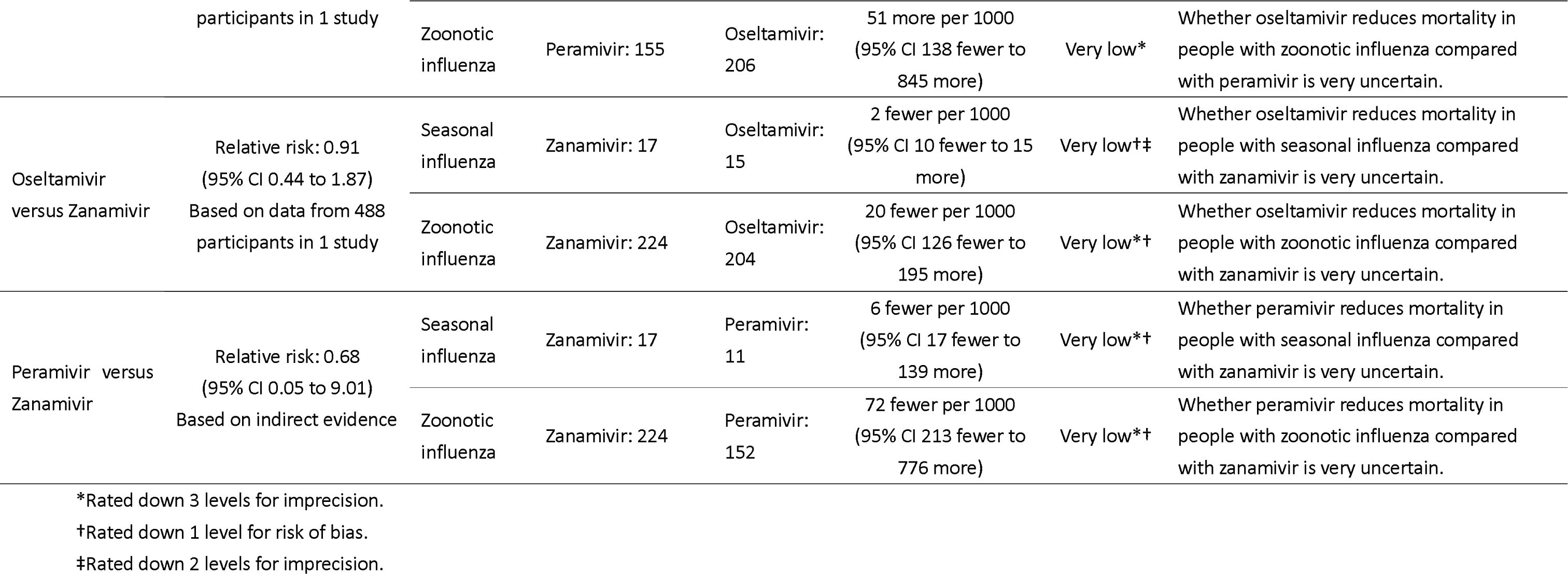
GRADE summary of findings for mortality for different comparisons.

**Table 3.** GRADE summary of findings for duration of hospitalization for different comparisons.

| Comparison | Mean difference<br>(95% CI) | Certainty in effect<br>estimates | Plain language summary |
| --- | --- | --- | --- |
| Oseltamivir versus<br>Standard care/placebo | -1.63 (-2.81 to<br>-0.45) | Low†‡ | Oseltamivir may reduce duration of hospitalization. |
| Peramivir versus<br>Standard care/placebo | -1.73 (-3.33 to<br>-0.13) | Low†‡ | Peramivir may reduce duration of hospitalization. |
| Oseltamivir versus<br>Peramivir | 0.10 (-0.98 to 1.18) | Low†‡ | There may be little or no difference between oseltamivir<br>and peramivir in duration of hospitalization. |

†Rated down 1 level for risk of bias.
‡Rated down 1 level for imprecision.

#### Mortality

The network meta-analysis of mortality included 4 trials of oseltamivir, peramivir or zanamivir involving 813 patients with severe seasonal influenza.^45,47,49,50^ The risk differences for the effect of oseltamivir, peramivir, or zanamivir on mortality when compared with placebo or standard care, or with each other varied from 18 fewer to 4 more per 1000 patients for seasonal influenza and from 232 fewer to 51 more per 1000 patients for zoonotic influenza (very low certainty evidence; Table 2). *Admission to ICU*

The network meta-analysis of ICU admission included two trials of oseltamivir or peramivir among 235 patients with severe seasonal influenza.^45,47^ The risk differences for the effect of oseltamivir or peramivir on ICU admission compared with standard care, or with each other varied from 29 fewer to 43 more per 1000 patients (Appendix 9.1).

#### Duration of hospitalization

The network meta-analysis of duration of hospitalization included three trials of oseltamivir or peramivir involving 226 patients with severe seasonal influenza.^44,47,50^ The MD in hospitalization duration for oseltamivir compared to placebo or standard care was 1.63 days shorter (95% CI −2.81 to −0.45 days; low certainty evidence). The MD for peramivir compared to placebo or standard care was 1.73 days shorter (95% CI −3.33 to −0.13 days; low certainty evidence). The MD in hospitalization duration for oseltamivir compared with peramivir was 0.10 days longer (95% CI −0.98 to 1.18 days; low certainty evidence; Table 3).

#### Time to alleviation of symptoms

The network meta-analysis of time to alleviation of symptoms included three trials with 283 patients with severe seasonal influenza that assessed the effect of oseltamivir or peramivir.^43,45,47^ The MD in time to alleviation of symptoms for oseltamivir compared to standard care was 0.34 days longer (95% CI −0.86 to 1.54 days; low certainty evidence). The MD in time to alleviation of symptoms for peramivir compared to standard care was 0.05 days shorter (95% CI −0.69 to 0.59 days; low certainty evidence; Appendix 9.2).

#### Adverse events

Two trials with 752 patients with severe influenza provided data on any adverse events and serious adverse events for comparing oseltamivir, peramivir, and zanamivir.^47,49^ There were no convincing differences in any adverse events or serious adverse events among oseltamivir, peramivir and zanamivir (very low certainty evidence; Appendix 9.3 and 9.4).

### Pairwise meta-analysis

Only one study^49^ reported data for patients with severe influenza on progression to invasive mechanical ventilation, duration of mechanical ventilation, emergence of resistance, and adverse events related to oseltamivir or zanamivir treatment. We were unable to conduct network meta-analyses for these outcomes but performed pairwise meta-analyses. Compared with zanamivir, the relative risks of oseltamivir on progression to mechanical ventilation, emergence of antiviral resistance, or adverse events related to treatment varied from 1.20 to 2.89 with 95% CIs overlapping with the null effect (very low certainty evidence; Appendix 9.5). The MD in duration of mechanical ventilation was 0.89 days (95% CI −2.32 to 4.10 days, very low certainty evidence; Appendix 9.6).

One study investigated combination treatment with baloxavir plus NAIs versus monotherapy with NAIs.^48^ There were few or no differences between baloxavir plus NAIs and NAIs alone on duration of hospitalization (MD 0.31 days shorter, 95% CI - 0.73 to 0.11 days; low certainty evidence) or emergence of antiviral resistance (RD 25 fewer per 1000, 95% CI 39 fewer to 42 more; low certainty evidence). Very low certainty evidence was available on the effects of baloxavir plus NAIs on ICU admission, mechanical ventilation, mortality, or adverse events compared to NAIs alone (Appendi× 10).

One study compared combination zanamivir plus rimantadine versus rimantadine alone.^46^ Very low certainty evidence was available on the effects of zanamivir plus rimantadine on duration of hospitalization, mortality, or adverse events compared with rimantadine monotherapy (Appendix 11).

### Additional analysis

Owing to the small number of eligible trials, we could not perform planned subgroup analyses or test for publication bias.

## Discussion

This network meta-analysis found that oseltamivir and peramivir may reduce duration of hospitalization in patients with severe seasonal influenza compared with placebo or standard care, but the evidence was of low certainty due to limited data from the small number of included RCTs. The effects of oseltamivir, peramivir, or zanamivir on mortality in patients with severe seasonal influenza or severe zoonotic influenza compared with placebo or standard care are very uncertain. Uncertainty also remains about the effects of oseltamivir, peramivir, and zanamivir on ICU admission. We did not find evidence of differences in any adverse events or serious adverse events among oseltamivir, peramivir, and zanamivir.

### Strengths and limitations

This is the first systematic review and network meta-analysis to evaluate the efficacy and safety of different antivirals for treatment of severe influenza. We focused on evidence for approved antivirals from RCTs, assessed the certainty of evidence using the GRADE approach, and presented absolute effects for outcomes. To reflect typical clinical scenarios in practice, we used two separate baseline risks for mortality and separately estimated absolute effects for severe seasonal influenza and zoonotic influenza. The selection of patient-important outcomes, baseline risks, and MID values for outcomes were based on the WHO guideline panel’s discussions and suggestions. The WHO panel also reviewed the results and assisted in their interpretation, ensuring a consistent interpretation of the available evidence to date. This systematic review provides the evidence base for the WHO clinical guideline recommendations for antiviral treatment of severe influenza.

Our review has limitations. First, only eight eligible trials were identified, and six trials were included in the network meta-analyses. Only one trial that compared oseltamivir to zanamivir provided data on progression to mechanical ventilation, duration of mechanical ventilation, emergence of resistance, and adverse events related to antiviral treatment.^49^ No trials addressed the effects of antivirals versus placebo or standard care on any adverse events or serious adverse events. Therefore, uncertainty remains about the effects of antivirals on most outcomes for patients with severe influenza. Second, due to sparse data available, we could not perform any prespecified subgroup analyses. Similarly, the assessments of incoherence and heterogeneity were not applicable for most outcomes and evaluation of publication bias was not applicable for all outcomes. Third, the WHO guideline panel suggested estimating separate absolute effects of antivirals on mortality for hospitalized patients with seasonal influenza and for zoonotic influenza. Since nearly all participants included in the eligible trials were patients with severe seasonal influenza, we estimated the absolute effects for patients with severe zoonotic influenza using the network relative estimates for severe seasonal influenza and baseline risk from a meta-analysis. Fourth, due to rating down the available evidence for risk of bias and imprecision, the certainty of evidence was low or very low for all available comparisons and outcomes. As new data from clinical trials become available,^51,52^ we anticipate that the certainty of evidence will improve for the outcomes of interest. To provide up-to-date evidence, we will periodically update this systematic review and network meta-analysis.

### Comparisons with other studies

One previous pairwise meta-analysis of 90 studies (all observational studies) of antiviral treatment of hospitalized patients with pandemic influenza A(H1N1)pdm09 virus infection reported that NAI treatment at any time versus none was associated with a nonsignificant reduction in mortality, but early NAI treatment (≤48 hours after symptom onset) versus late, and early antiviral treatment initiation versus none, was associated with significant reductions in mortality.^15^ Another individual participant data meta-analysis that included 29234 hospitalized patients with pandemic influenza A(H1N1pdm09) virus infection from 78 observational studies, reported that NAI treatment (irrespective of timing) was associated with a reduction in mortality compared with no treatment, and early treatment (within 2 days of symptom onset) was associated with a reduction in mortality relative to later treatment or no treatment.^16^ These meta-analyses reported inconsistent results regarding NAI treatment of patients with severe influenza at any time versus no NAI treatment on mortality, mainly because of the different kinds of data (aggregate data versus individual participant data) used.

Our network meta-analysis, including only RCTs, did not substantiate the findings of prior meta-analyses. We assessed the effect of each antiviral on patient-important outcomes and presented absolute effects for mortality in severe seasonal influenza patients and estimated absolute effects for mortality in severe zoonotic influenza patients, although the very low certainty evidence indicated low confidence in inferences regarding mortality. Moreover, since all included trials did not present data related to the timing of antiviral treatment initiation in relationship to symptom onset, we were unable to examine the effect of the timing of antiviral treatment initiation from symptom onset on outcomes.

One pairwise meta-analysis that included 7 RCTs, addressed different dosages and regimens of NAIs in hospitalized patients with seasonal or pandemic influenza and reported non-significant differences among different antiviral treatment regimens on mortality, time to clinical resolution, and viral clearance.^18^ These findings regarding time to clinical resolution are consistent with our results of antivirals not having important effects on reducing time to alleviation of symptoms in patients with severe influenza.

One previous individual participant data meta-analysis that included observational studies of patients hospitalized with pandemic influenza A(H1N1)pdm09 virus infection, reported that NAI treatment started on the day of admission, regardless of time since symptom onset, was associated with a reduction in the length of hospital stay compared with no or later initiation of NAI treatment.^19^ Our meta-analysis also found that oseltamivir and peramivir may reduce the duration of hospitalization in patients with severe influenza compared with placebo or standard care. The WHO guideline panel discussed the evidence from observational studies and deemed that they did not provide a higher certainty of evidence for this population compared to the current systematic review of RCTs.

### Implications for future research and practice

Due to limited data from the small number of RCTs of antivirals for treatment of patients with severe seasonal influenza and lack of RCTs for treatment of severe zoonotic influenza, the current level of evidence for antiviral treatment of severe seasonal or zoonotic influenza is of low certainty. Additional clinical trials of antivirals are needed to inform the clinical benefit, safety, and effects on antiviral resistance in patients with severe influenza. Important gaps include better evidence of the effects of antiviral treatment of patients with severe influenza on admission to ICU, progression to invasive mechanical ventilation, duration of mechanical ventilation, mortality, and emergence of antiviral resistance, and to evaluate the effects of antivirals on outcomes in important subgroup populations, including patients with severe zoonotic influenza.

## Conclusions

Data from RCTs of antiviral treatment of patients with severe influenza are limited. In patients with severe influenza, oseltamivir or peramivir may reduce the duration of hospitalization compared with placebo or standard care. There is high uncertainty of the effects of oseltamivir, peramivir, and zanamivir in patients with severe seasonal or zoonotic influenza on ICU admission and mortality. Sufficiently powered clinical trials in patients with severe influenza due to seasonal influenza virus infections and novel influenza A virus infections are needed to provide higher certainty evidence of the effects of antiviral treatment on important clinical outcomes.

## Supporting information

PRISMA

## Data Availability

All data produced in the present work are contained in the manuscript

## Contributors

GG, QH, and YG conceived and designed the study. YG and QH designed and performed the search strategy. YG, ML, YZ, SL, and XC screened and selected the articles. YG, ML, YC, YS, JX, QZ, ZL, and WZ extracted the data and assessed the risk of bias. YG and QH analyzed the data. GG and JT supervised the data analyses. YG and QH rated the certainty of evidence. GG provided methodological advice on baseline risk selection and GRADE assessment. YG, GG, TU, and QH interpreted the data. YG and QH drafted the manuscript. YG, GG, TU, and QH revised the manuscript. All authors approved the final version of the manuscript. YG and QH accessed and verified the underlying data. All authors had full access to all the data in the study and had final responsibility for the decision to submit for publication.

## Data sharing statement

Data in this systematic review with meta-analysis are extracted from published studies available on the internet. All processed data are presented in this article and the appendix.

## Acknowledgements

We thank members of the WHO for critical feedback on the review question, subgroup and outcome selection, and GRADE judgments: Janet Diaz (World Health Organization, Geneva, Switzerland); Steven Mcgloughlin (World Health Organization, Geneva, Switzerland); Jamie Rylance (World Health Organization, Geneva, Switzerland). We thank Rachel Couban (librarian at McMaster University) for helping with developing the search strategy. We thank Ping Liu (The First Affiliated Hospital of Chongqing Medical University, Chongqing, China) for contributing to the literature screening.

## Disclaimer

The findings and conclusions in this report are those of the authors and do not necessarily represent the official position of the Centers for Disease Control and Prevention.

