## Appendix for "Antivirals for treatment of severe influenza: a systematic review and network meta-analysis of randomized controlled trials"

### Appendix 1. Search strategy for databases

**Ovid MEDLINE(R) ALL**

1	exp Influenza, Human/

2	exp Influenza A virus/

3	exp Influenza B virus/

4	exp Influenzavirus C/

5	(Influenza or flu or H1N1 or PH1N1 or H3N2 or AH1N1 or AH3N2 or H5N1 or H7N9).mp. [mp=title, book title, abstract, original title, name of substance word, subject heading word, floating sub-heading word, keyword heading word, organism supplementary concept word, protocol supplementary concept word, rare disease supplementary concept word, unique identifier, synonyms, population supplementary concept word, anatomy supplementary concept word]

6	or/1-5

7	Antiviral agents/

8	Antiviral*.tw.

9	(neuraminidase inhibitor* or NA inhibitor*).tw.

10	Oseltamivir/ or Zanamivir/

11	(oseltamivir or tamiflu or "GS 4104" or GS4104 or GS-4104 or "GS 4071" or GS4071 or GS-4071 or zanamivir or relenza or "GG 167" or GG167 or GG-167 or CS-8958 or Dectova or Laninamivir or R-125489 or R125489 or "R 125489" or Inavir or peramivir or "BCX 1812" or BCX1812 or BCX-1812 or "RWJ 270201" or RWJ270201 or RWJ-270201 or Rapivab or rapiacta).ti,ab.

12	Viral Polymerase Complex Inhibitor*.tw.

13	(Favipiravir or T-705 or Avigan or FabiFlu or Pimodivir or VX-787 or JNJ-63623872 or AL-794 or ALS-033719 or ZSP1273 or Enisamium iodide or FAV00A or TG-1000 or GP681).ti,ab.

14	matrix protein 2 ion channel inhibitor*.tw.

15	(Radavirsen or AVI-7100).ti,ab.

16	cap-dependent endonuclease inhibitor*.tw.

17	("Baloxavir marboxil" or Baloxavir or S-033188 or Xofluza).ti,ab.

18	(Umifenovir or Arbidol or Arbidole).ti,ab.

19	Amantadine/ or Rimantadine/

20	(Amantadine or Symmetrel or Symetrel or Rimantadine or Flumadine or Roflual).ti,ab.

21	or/7-20

22	6 and 21

23	randomized controlled trial.pt.

24	controlled clinical trial.pt.

25	randomized.ab.

26	placebo.ab.

27	drug therapy.fs.

28	randomly.ab.

29	trial.ti.

30	groups.ab.

31	or/23-30

32	(animals not (humans and animals)).sh.

33	31 not 32

34	22 and 33

**Ovid Embase**

1	exp Influenza/ or Influenza virus/

2	exp Influenza A virus/ or exp Influenza A virus/

3	exp Influenza B/ or exp Influenza B virus/

4	exp Influenza C/ or exp Influenza C virus/

5	(Influenza or flu or H1N1 or PH1N1 or H3N2 or AH1N1 or AH3N2 or H5N1 or H7N9).mp. [mp=title, abstract, heading word, drug trade name, original title, device manufacturer, drug manufacturer, device trade name, keyword heading word, floating subheading word, candidate term word]

6	or/1-5

7	Antivirus agent/

8	Antiviral*.tw.

9	(neuraminidase inhibitor* or NA inhibitor*).tw.

10	Sialidase inhibitor/ or Oseltamivir/ or Zanamivir/ or Laninamivir/ or Peramivir/

11	(oseltamivir or tamiflu or "GS 4104" or GS4104 or GS-4104 or "GS 4071" or GS4071 or GS-4071 or zanamivir or relenza or "GG 167" or GG167 or GG-167 or CS-8958 or Dectova or Laninamivir or R-125489 or R125489 or "R 125489" or Inavir or peramivir or "BCX 1812" or BCX1812 or BCX-1812 or "RWJ 270201" or RWJ270201 or RWJ-270201 or Rapivab or rapiacta).ti,ab.

12	Viral Polymerase Complex Inhibitor*.tw.

13	Favipiravir/ or Pimodivir/ or (Favipiravir or T-705 or Avigan or FabiFlu or Pimodivir or VX-787 or JNJ-63623872 or AL-794 or ALS-033719 or ZSP1273 or Enisamium iodide or FAV00A or TG-1000 or GP681).ti,ab.

14	matrix protein 2 ion channel inhibitor*.tw.

15	Radavirsen/ or (Radavirsen or AVI-7100).ti,ab.

16	cap-dependent endonuclease inhibitor*.tw.

17	Baloxavir marboxil/ or ("Baloxavir marboxil" or Baloxavir or S-033188 or Xofluza).ti,ab.

18	Umifenovir/ or (Umifenovir or Arbidol or Arbidole).ti,ab.

19	Amantadine/ or Rimantadine/

20	(Amantadine or Symmetrel or Symetrel or Rimantadine or Flumadine or Roflual).ti,ab.

21	or/7-20

22	6 and 21

23	Randomized controlled trial/

24	Controlled clinical study/

25	random$.ti,ab.

26	randomization/

27	intermethod comparison/

28	placebo.ti,ab.

29	(compare or compared or comparison).ti.

30	((evaluated or evaluate or evaluating or assessed or assess) and (compare or compared or comparing or comparison)).ab.

31	(open adj label).ti,ab.

32	((double or single or doubly or singly) adj (blind or blinded or blindly)).ti,ab.

33	double blind procedure/

34	parallel group$1.ti,ab.

35	(crossover or cross over).ti,ab.

36	((assign$ or match or matched or allocation) adj5 (alternate or group$1 or intervention$1 or patient$1 or subject$1 or participant$1)).ti,ab.

37	(assigned or allocated).ti,ab.

38	(controlled adj7 (study or design or trial)).ti,ab.

39	(volunteer or volunteers).ti,ab.

40	human experiment/

41	trial.ti.

42	or/23-41

43	(random$ adj sampl$ adj7 ("cross section$" or questionnaire$1 or survey$ or database$1)).ti,ab. not (comparative study/ or controlled study/ or randomi?ed controlled.ti,ab. or randomly assigned.ti,ab.)

44	Cross-sectional study/ not (randomized controlled trial/ or controlled clinical study/ or controlled study/ or randomi?ed controlled.ti,ab. or control group$1.ti,ab.)

45	(((case adj control$) and random$) not randomi?ed controlled).ti,ab.

46	(Systematic review not (trial or study)).ti.

47	(nonrandom$ not random$).ti,ab.

48	"Random field$".ti,ab.

49	(random cluster adj3 sampl$).ti,ab.

50	(review.ab. and review.pt.) not trial.ti.

51	"we searched".ab. and (review.ti. or review.pt.)

52	"update review".ab.

53	(databases adj4 searched).ab.

54	(rat or rats or mouse or mice or swine or porcine or murine or sheep or lambs or pigs or piglets or rabbit or rabbits or cat or cats or dog or dogs or cattle or bovine or monkey or monkeys or trout or marmoset$1).ti. and animal experiment/

55	Animal experiment/ not (human experiment/ or human/)

56	or/43-55

57	42 not 56

58	22 and 57

**Cochrane Central Register of Controlled Trials**

1	exp Influenza, Human/

2	exp Influenza A virus/

3	exp Influenza B virus/

4	exp Influenzavirus C/

5	(Influenza or flu or H1N1 or PH1N1 or H3N2 or AH1N1 or AH3N2 or H5N1 or H7N9).mp. [mp=title, original title, abstract, floating sub-heading word, mesh headings, heading words, keyword]

6	or/1-5

7	Antiviral agents/

8	Antiviral*.tw.

9	(neuraminidase inhibitor* or NA inhibitor*).tw.

10	Oseltamivir/ or Zanamivir/

11	(oseltamivir or tamiflu or "GS 4104" or GS4104 or GS-4104 or "GS 4071" or GS4071 or GS-4071 or zanamivir or relenza or "GG 167" or GG167 or GG-167 or CS-8958 or Dectova or Laninamivir or R-125489 or R125489 or "R 125489" or Inavir or peramivir or "BCX 1812" or BCX1812 or BCX-1812 or "RWJ 270201" or RWJ270201 or RWJ-270201 or Rapivab or rapiacta).ti,ab.

12	Viral Polymerase Complex Inhibitor*.tw.

13	(Favipiravir or T-705 or Avigan or FabiFlu or Pimodivir or VX-787 or JNJ-63623872 or AL-794 or ALS-033719 or ZSP1273 or Enisamium iodide or FAV00A or TG-1000 or GP681).ti,ab.

14	matrix protein 2 ion channel inhibitor*.tw.

15	(Radavirsen or AVI-7100).ti,ab.

16	cap-dependent endonuclease inhibitor*.tw.

17	("Baloxavir marboxil" or Baloxavir or S-033188 or Xofluza).ti,ab.

18	(Umifenovir or Arbidol or Arbidole).ti,ab.

19	Amantadine/ or Rimantadine/

20	(Amantadine or Symmetrel or Symetrel or Rimantadine or Flumadine or Roflual).ti,ab.

21	or/7-20

22	6 and 21

23	randomized controlled trial.pt.

24	controlled clinical trial.pt.

25	randomized.ab.

26	placebo.ab.

27	drug therapy.fs.

28	randomly.ab.

29	trial.ti.

30	groups.ab.

31	or/23-30

32	(animals not (humans and animals)).sh.

33	31 not 32

34	22 and 33

**Global Health**

1	exp Influenza/ or Influenza viruses/

2	exp Influenza A virus/ or exp Influenza A virus/

3	exp Influenza B/ or exp Influenza B virus/

4	exp Influenza C/ or exp Influenza C virus/

5	(Influenza or flu or H1N1 or PH1N1 or H3N2 or AH1N1 or AH3N2 or H5N1 or H7N9).mp. [mp=abstract, title, original title, heading words, cabicodes words]

6	or/1-5

7	Antiviral agents/

8	Antiviral*.tw.

9	(neuraminidase inhibitor* or NA inhibitor*).tw.

10	Sialidase inhibitors/ or Oseltamivir/ or Zanamivir/ or Laninamivir/ or Peramivir/

11	(oseltamivir or tamiflu or "GS 4104" or GS4104 or GS-4104 or "GS 4071" or GS4071 or GS-4071 or zanamivir or relenza or "GG 167" or GG167 or GG-167 or CS-8958 or Dectova or Laninamivir or R-125489 or R125489 or "R 125489" or Inavir or peramivir or "BCX 1812" or BCX1812 or BCX-1812 or "RWJ 270201" or RWJ270201 or RWJ-270201 or Rapivab or rapiacta).ti,ab.

12	Viral Polymerase Complex Inhibitor*.tw.

13	Favipiravir/ or (Favipiravir or T-705 or Avigan or FabiFlu or Pimodivir or VX-787 or JNJ-63623872 or AL-794 or ALS-033719 or ZSP1273 or Enisamium iodide or FAV00A or TG-1000 or GP681).ti,ab.

14	matrix protein 2 ion channel inhibitor*.tw.

15	(Radavirsen or AVI-7100).ti,ab.

16	cap-dependent endonuclease inhibitor*.tw.

17	("Baloxavir marboxil" or Baloxavir or S-033188 or Xofluza).ti,ab.

18	(Umifenovir or Arbidol or Arbidole).ti,ab.

19	Amantadine/ or Rimantadine/

20	(Amantadine or Symmetrel or Symetrel or Rimantadine or Flumadine or Roflual).ti,ab.

21	or/7-20

22	6 and 21

23	exp randomized controlled trials/

24	(randomized controlled trial or random* or blind* or placebo*).mp. [mp=abstract, title, original title, heading words, cabicodes words]

25	23 or 24

26	22 and 25

**CINAHL**

| **#** | **Query** |
| --- | --- |
| S36 | S23 AND S26 AND S35 |
| S35 | S27 OR S28 OR S29 OR S30 OR S31 OR S32 OR S33 OR S34 |
| S34 | TI ( matrix protein 2 ion channel inhibitor* OR Radavirsen or AVI-7100 OR cap-dependent endonuclease inhibitor* OR "Baloxavir marboxil" or Baloxavir or S-033188 or Xofluza OR Umifenovir or Arbidol or Arbidole OR Amantadine or Symmetrel or Symetrel or Rimantadine or Flumadine or Roflual ) OR AB ( matrix protein 2 ion channel inhibitor* OR Radavirsen or AVI-7100 OR cap-dependent endonuclease inhibitor* OR "Baloxavir marboxil" or Baloxavir or S-033188 or Xofluza OR Umifenovir or Arbidol or Arbidole OR Amantadine or Symmetrel or Symetrel or Rimantadine or Flumadine or Roflual ) |
| S33 | (MH "Amantadine") |
| S32 | TI ( Viral Polymerase Complex Inhibitor* OR Favipiravir or T-705 or Avigan or FabiFlu or Pimodivir or VX-787 or JNJ-63623872 or AL-794 or ALS-033719 or ZSP1273 or Enisamium iodide or FAV00A or TG-1000 or GP681 ) OR AB ( Viral Polymerase Complex Inhibitor* OR Favipiravir or T-705 or Avigan or FabiFlu or Pimodivir or VX-787 or JNJ-63623872 or AL-794 or ALS-033719 or ZSP1273 or Enisamium iodide or FAV00A or TG-1000 or GP681 ) |
| S31 | TI ( oseltamivir or tamiflu or "GS 4104" or GS4104 or GS-4104 or "GS 4071" or GS4071 or GS-4071 or zanamivir or relenza or "GG 167" or GG167 or GG-167 or CS-8958 or Dectova or Laninamivir or R-125489 or R125489 or "R 125489" or Inavir or peramivir or "BCX 1812" or BCX1812 or BCX-1812 or "RWJ 270201" or RWJ270201 or RWJ-270201 or Rapivab or rapiacta ) OR AB ( oseltamivir or tamiflu or "GS 4104" or GS4104 or GS-4104 or "GS 4071" or GS4071 or GS-4071 or zanamivir or relenza or "GG 167" or GG167 or GG-167 or CS-8958 or Dectova or Laninamivir or R-125489 or R125489 or "R 125489" or Inavir or peramivir or "BCX 1812" or BCX1812 or BCX-1812 or "RWJ 270201" or RWJ270201 or RWJ-270201 or Rapivab or rapiacta ) |
| S30 | (MH "Oseltamivir") |
| S29 | TI(neuraminidase inhibitor* or NA inhibitor*) OR AB(neuraminidase inhibitor* or NA inhibitor*) |
| S28 | TI Antiviral* OR AB Antiviral* |
| S27 | (MH "Antiviral Agents") |
| S26 | S24 OR S25 |
| S25 | TI ( Influenza or flu or H1N1 or PH1N1 or H3N2 or AH1N1 or AH3N2 or H5N1 or H7N9 ) OR AB ( Influenza or flu or H1N1 or PH1N1 or H3N2 or AH1N1 or AH3N2 or H5N1 or H7N9 ) |
| S24 | (MH "Influenza+") OR (MH "Influenza A Virus+") OR (MH "Influenzavirus C") OR (MH "Influenza B Virus") |
| S23 | S22 NOT S21 |
| S22 | S1 OR S2 OR S3 OR S4 OR S5 OR S6 OR S7 OR S8 OR S9 OR S10 OR S11 OR S12 OR S13 OR S14 OR S15 |
| S21 | S19 NOT S20 |
| S20 | MH (human) |
| S19 | S16 OR S17 OR S18 |
| S18 | TI (animal model*) |
| S17 | MH (animal studies) |
| S16 | MH animals+ |
| S15 | AB (cluster W3 RCT) |
| S14 | MH (crossover design) OR MH (comparative studies) |
| S13 | AB (control W5 group) |
| S12 | PT (randomized controlled trial) |
| S11 | MH (placebos) |
| S10 | MH (sample size) AND AB (assigned OR allocated OR control) |
| S9 | TI (trial) |
| S8 | AB (random*) |
| S7 | TI (randomised OR randomized) |
| S6 | MH cluster sample |
| S5 | MH pretest‐posttest design |
| S4 | MH random assignment |
| S3 | MH single‐blind studies |
| S2 | MH double‐blind studies |
| S1 | MH randomized controlled trials |

**Epistemonikos**

Influenza antivirals

**ClinIcaltrial.gov**

Influenza antivirals

### Appendix 2. Details of methods

#### 2.1. Details of data extraction

Pairs of reviewers independently extracted the following data: study characteristics (first author, trial registration, publication year, publication status, country, and sample size); participant characteristics (age, sex, disease severity, comorbidities, influenza virus type); characteristics of antivirals (dosing, frequency, route of administration, treatment duration, and length of follow-up); and outcomes. Reviewers resolved discrepancies by discussion or, if necessary, through consultation with a third reviewer.

#### 2.2. Details of risk of bias assessment

To evaluate the risk of bias of eligible RCTs, we used a modified Cochrane risk of bias tool, including assessing the following domains: random sequence generation; allocation concealment; blinding of participants, healthcare providers, data collectors, outcome assessor/adjudicator, and data analysts; incomplete outcome data (≥ 10% missing data was considered high risk of bias); selective outcome reporting; and other sources of bias (i.e. baseline imbalance, early trial discontinuation). Pairs of reviewers independently rated each domain at the outcome level as: high, probably high, probably low, or low risk of bias. Because lack of blinding is unlikely to bias assessment of mortality, admission to ICU, progression to invasive mechanical ventilation, and emergence of antiviral resistance, we rated the blinding for these outcomes as low risk of bias, regardless of blinding status. Reviewers resolved discrepancies by discussion or, if necessary, with adjudication by a third party.

### Appendix 3. Additional characteristics of eligible RCTs

| **Study** | **Pregnant %** | **Inpatient %** | **Intensive care %** | **Patients received influenza vaccination %** | **Outcomes** |
| --- | --- | --- | --- | --- | --- |
| Chen 2021 | NR | 100 | NR | 0 | Time to alleviation of symptoms |
| Dawood 2016 | 0 | 100 | 3.33 | NR | Duration of hospitalization |
| de Jong 2014 | 0 | 100 | 19.01 | 4.96 | Mortality  Admission to ICU  Time to alleviation of symptoms |
| Ison 2003 | 0 | 100 | 14.63 | NR | Mortality  Any adverse events  Serious adverse events  Duration of hospitalization |
| Ison 2013 | 0 | 100 | NR | NR | Mortality  Admission to ICU  Any adverse events  Serious adverse events  Duration of hospitalization  Time to alleviation of symptoms |
| Kumar 2022 | 0 | 100 | 13.55 | NR | Mortality  Admission to ICU  Progression to mechanical ventilation  Emergence of resistance  Adverse events related to antivirals  Any adverse events  Serious adverse events  Duration of mechanical ventilation  Duration of hospitalization |
| Marty 2017 | 0 | 100 | 39.67 | 10.89 | Mortality  Progression to mechanical ventilation  Emergence of resistance  Adverse events related to antivirals  Any adverse events  Serious adverse events  Duration of mechanical ventilation |
| Ramirez 2018 | 0 | 100 | NR | NR | Mortality  Duration of hospitalization |

### Appendix 4. Risk of bias for eligible studies

| **Study** | **Sequence generation** | **Allocation concealment** | **Blinding of patients** | **Blinding of health care providers** | **Blinding of data collectors** | **Blinding of outcome assessors/ adjudicators** | **Blinding of data analysts** | **Incomplete outcome data** | **Selective outcome reporting** | **Other bias** |
| --- | --- | --- | --- | --- | --- | --- | --- | --- | --- | --- |
| **Mortality** | | | | | | | | | | |
| de Jong 2014 | Probably Low | Probably High | Low | Low | Low | Low | Low | Low | Probably Low | Low |
| Ison 2003 | Low | Low | Low | Low | Low | Low | Low | High | Probably Low | Probably High |
| Ison 2013 | Probably Low | Low | Low | Low | Low | Low | Low | Low | Low | Low |
| Kumar 2022 | Low | Low | Low | Low | Low | Low | Low | Low | Low | Low |
| Marty 2017 | Low | Low | Low | Low | Low | Low | Low | High | Low | Low |
| Ramirez 2018 | Low | Probably Low | Low | Low | Low | Low | Low | Low | Low | Low |
| **Admission to ICU** | | | | | | | | | | |
| de Jong 2014 | Probably Low | Probably High | Low | Low | Low | Low | Low | Low | Low | Low |
| Ison 2013 | Probably Low | Low | Low | Low | Low | Low | Low | Low | Low | Low |
| Kumar 2022 | Low | Low | Low | Low | Low | Low | Low | Low | Low | Low |
| **Progression to mechanical ventilation** | | | | | | | | | | |
| Kumar 2022 | Low | Low | Low | Low | Low | Low | Low | Low | Probably Low | Low |
| Marty 2017 | Low | Low | Low | Low | Low | Low | Low | High | Probably Low | Low |
| **Emergence of resistance** | | | | | | | | | | |
| Kumar 2022 | Low | Low | Low | Low | Low | Low | Low | Low | Low | Low |
| Marty 2017 | Low | Low | Low | Low | Low | Low | Low | High | Low | Low |
| **Any adverse events** | | | | | | | | | | |
| Ison 2003 | Low | Low | Low | Low | Probably High | Probably High | Probably High | High | Probably Low | Probably High |
| Ison 2013 | Probably Low | Low | Low | Low | Probably High | Probably High | Low | Low | Low | Low |
| Kumar 2022 | Low | Low | Low | Low | Low | Low | Low | Low | Low | Low |
| Marty 2017 | Low | Low | Low | Low | Probably High | Low | Probably High | High | Low | Low |
| **Adverse events related to treatments** | | | | | | | | | | |
| Kumar 2022 | Low | Low | Low | Low | Low | Low | Low | Low | Low | Low |
| Marty 2017 | Low | Low | Low | Low | Probably High | Low | Probably High | High | Low | Low |
| **Serious adverse events** | | | | | | | | | | |
| Ison 2003 | Low | Low | Low | Low | Probably High | Probably High | Probably High | High | Probably Low | Probably High |
| Ison 2013 | Probably Low | Low | Low | Low | Probably High | Probably High | Low | Low | Low | Low |
| Kumar 2022 | Low | Low | Low | Low | Low | Low | Low | Low | Low | Low |
| Marty 2017 | Low | Low | Low | Low | Probably High | Low | Probably High | High | Low | Low |
| **Duration of hospitalization** | | | | | | | | | | |
| Dawood 2016 | Low | Low | Low | Low | Probably High | Probably High | Probably High | Low | Low | Probably High |
| Ison 2003 | Low | Low | Low | Low | Probably High | Probably High | Probably High | High | Probably Low | Probably High |
| Ison 2013 | Probably Low | Low | Low | Low | Probably High | Probably High | Low | Low | Low | Low |
| Kumar 2022 | Low | Low | Low | Low | Low | Low | Low | Low | Low | Low |
| Ramirez 2018 | Low | Probably Low | High | High | High | High | High | Low | Low | Low |
| **Time to alleviation of symptoms** | | | | | | | | | | |
| Chen 2021 | Low | Probably High | Probably High | Probably High | Probably High | Probably High | Probably High | Low | Low | Low |
| de Jong 2014 | Probably Low | Probably High | Low | Low | Probably Low | Probably Low | Probably High | Low | Low | Low |
| Ison 2013 | Probably Low | Low | Low | Low | Probably High | Probably High | Low | Low | Low | Low |
| **Duration of mechanical ventilation** | | | | | | | | | | |
| Kumar 2022 | Low | Low | Low | Low | Low | Low | Low | Low | Low | Low |
| Marty 2017 | Low | Low | Low | Low | Probably High | Low | Low | High | Probably Low | Low |

### Appendix 5. Network plots

*The size of the circle represents the number of participants. The width of the line represents the number of studies.

#### 5.1. Network plot for admission to ICU

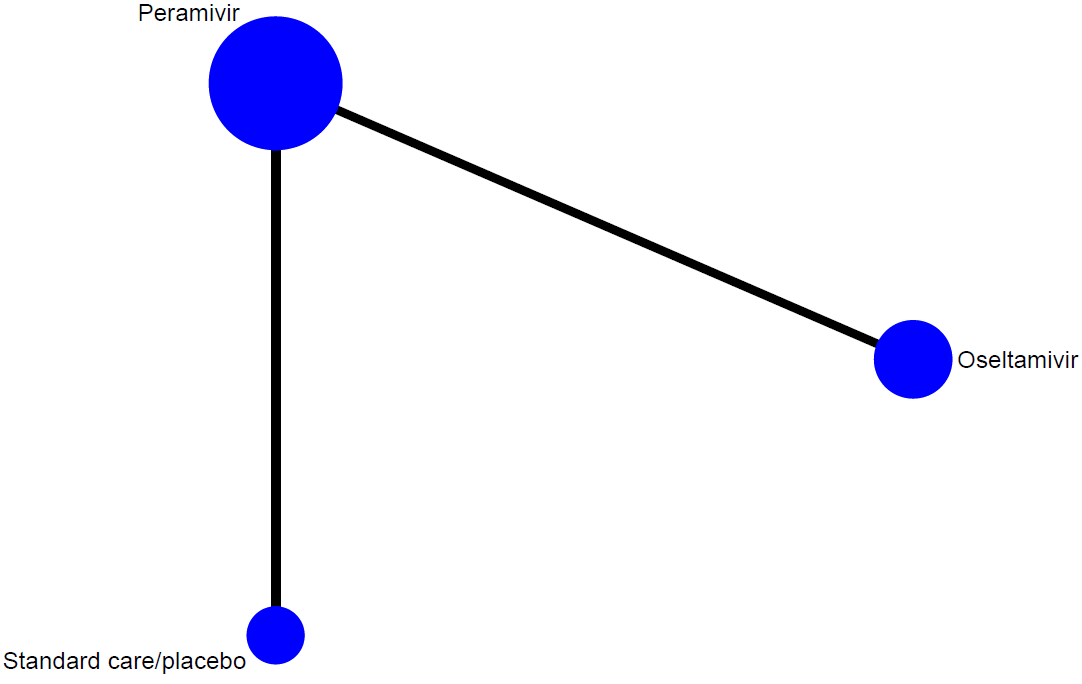

#### 5.2. Network plot for duration of hospitalization

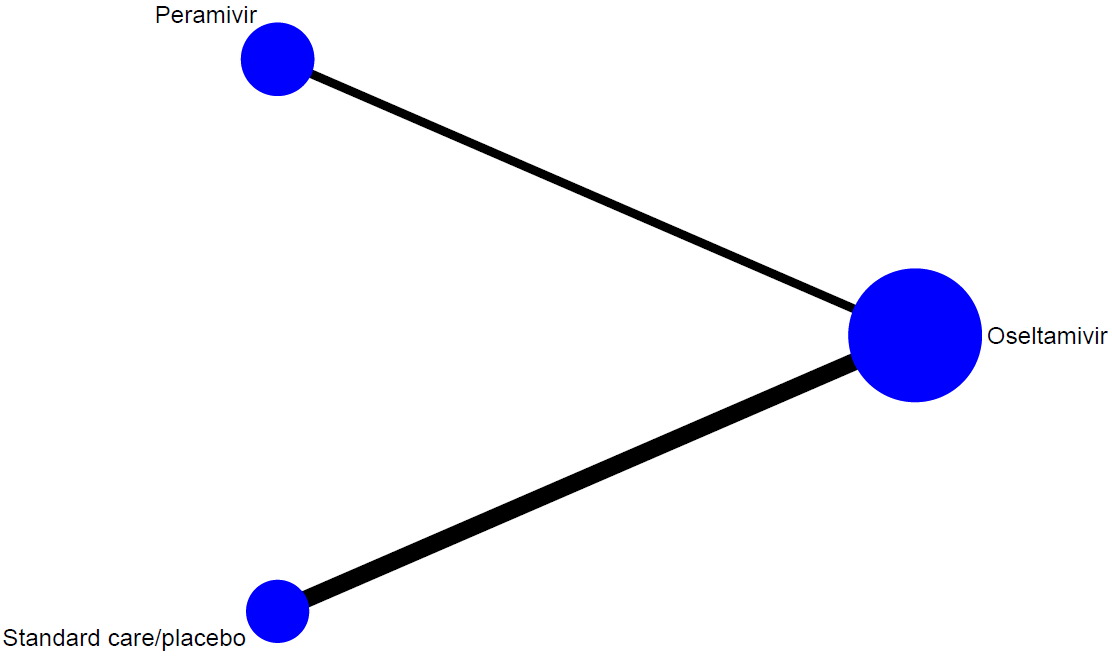

#### 5.3. Network plot for time to alleviation of symptoms

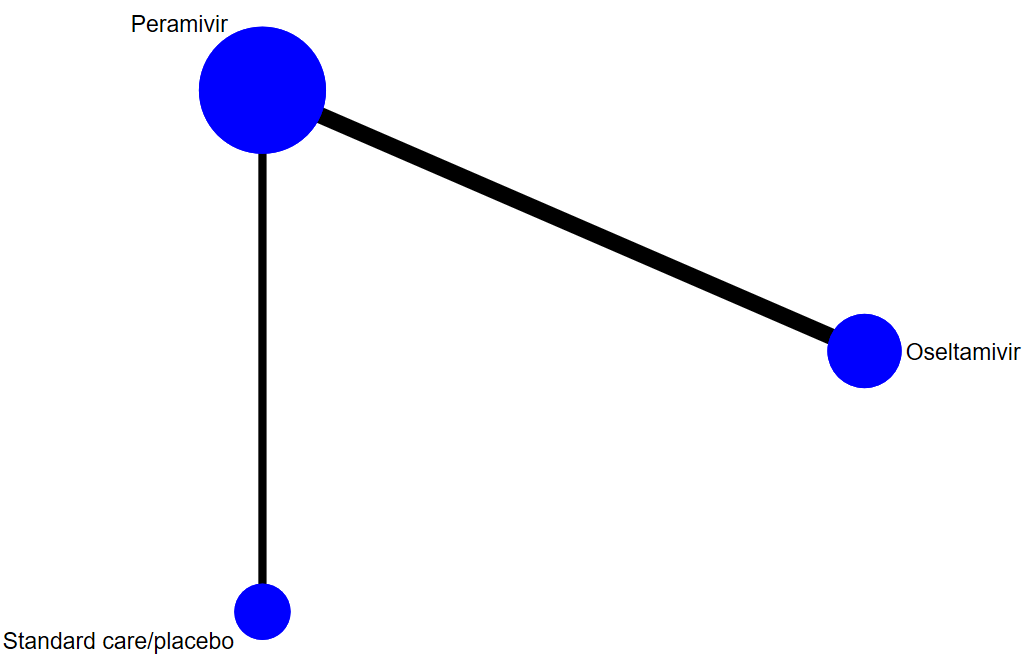

#### 5.4. Network plot for any adverse events

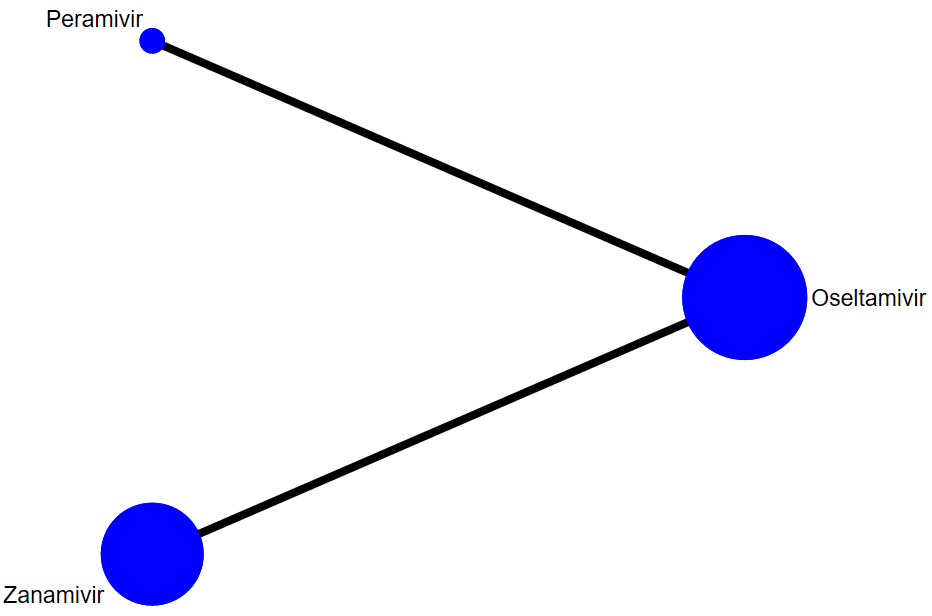

#### 5.5. Network plot for serious adverse events

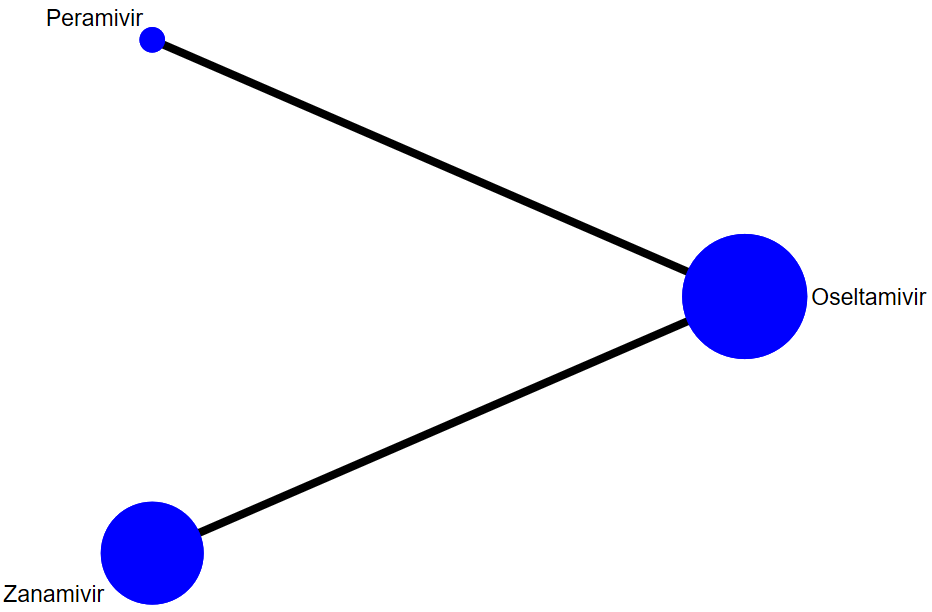

### Appendix 6. Assessment of between-study heterogeneity

| **Outcome** | **Comparison** | **No. study** | **I^2^** |
| --- | --- | --- | --- |
| Mortality | Oseltamivir vs. Peramivir | 1 | NA |
|  | Oseltamivir vs. Standard care/placebo | 1 | NA |
|  | Oseltamivir vs. Zanamivir | 1 | NA |
|  | Peramivir vs. Standard care/placebo | 1 | NA |
| Admission to ICU | Oseltamivir vs. Peramivir | 1 | NA |
|  | Peramivir vs. Standard care/placebo | 1 | NA |
| Time to alleviation of symptoms | Oseltamivir vs. Peramivir | 2 | 0% |
|  | Peramivir vs. Standard care/placebo | 1 | NA |
| Duration of hospitalization | Oseltamivir vs. Standard care/placebo | 2 | 0% |
|  | Oseltamivir vs. Peramivir | 1 | NA |
| Any adverse events | Oseltamivir vs. Peramivir | 1 | NA |
|  | Oseltamivir vs. Zanamivir | 1 | NA |
| Serious adverse events | Oseltamivir vs. Peramivir | 1 | NA |
|  | Oseltamivir vs. Zanamivir | 1 | NA |

NA, not applicable.

### Appendix 7. Assessment of global inconsistency

| **Outcome** | **P value** |
| --- | --- |
| Mortality | 0.487 |
| Admission to ICU | NA |
| Time to alleviation of symptoms | NA |
| Duration of hospitalization | NA |
| Any adverse events | NA |
| Serious adverse events | NA |

NA, not applicable.

### Appendix 8. Direct, indirect, and network treatment estimates

#### 8.1. Direct, indirect, and network treatment estimates for mortality

| **Comparison** | **k** | **Prop** | **NMA (95% CI)** | **Direct (95% CI)** | **Indirect (95% CI)** | **RoR (95% CI)** | **z** | **Incoherence p-value** |
| --- | --- | --- | --- | --- | --- | --- | --- | --- |
| Oseltamivir vs. Peramivir | 1 | 0.61 | 1.33 (0.11 to 15.87) | 0.66 (0.03 to 15.79) | 3.97 (0.08 to 207.32) | 0.17 (0.00 to 26.45) | -0.7 | 0.487 |
| Oseltamivir vs. Standard care/placebo | 1 | 0.78 | 0.53 (0.07 to 4.24) | 0.78 (0.07 to 8.17) | 0.13 (0.00 to 11.50) | 6.05 (0.04 to 969.21) | 0.7 | 0.487 |
| Oseltamivir vs. Zanamivir | 1 | 1 | 0.91 (0.44 to 1.87) | 0.91 (0.44 to 1.87) | NA | NA | NA | NA |
| Peramivir vs. Standard care/placebo | 1 | 0.61 | 0.40 (0.03 to 4.72) | 0.20 (0.01 to 4.69) | 1.18 (0.02 to 61.91) | 0.17 (0.00 to 26.45) | -0.7 | 0.487 |
| Peramivir vs. Zanamivir | 0 | 0 | 0.68 (0.05 to 9.01) | NA | 0.68 (0.05 to 9.01) | NA | NA | NA |
| Zanamivir vs. Standard care/placebo | 0 | 0 | 0.58 (0.06 to 5.29) | NA | 0.58 (0.06 to 5.29) | NA | NA | NA |

Comparison: treatment comparison; k: number of studies providing direct evidence; prop: direct evidence proportion; NMA: estimated treatment effect (RR) in network meta-analysis; direct: estimated treatment effect (RR) derived from direct evidence; indirect: estimated treatment effect (RR) derived from indirect evidence; RoR: Ratio of Ratios (direct versus indirect); z: z-value of test for disagreement (direct versus indirect): Incoherence p-value: p-value of test for disagreement (direct versus indirect). NA: not applicable.

#### 8.2. Direct, indirect, and network treatment estimates for admission to ICU

| **Comparison** | **k** | **Prop** | **NMA (95% CI)** | **Direct (95% CI)** | **Indirect (95% CI)** | **Diff (95% CI)** | **z** | **Incoherence p-value** |
| --- | --- | --- | --- | --- | --- | --- | --- | --- |
| Oseltamivir vs. Peramivir | 1 | 1 | 0.043 (-0.034 to 0.121) | 0.043 (-0.034 to 0.121) | NA | NA | NA | NA |
| Oseltamivir vs. Standard care/placebo | 0 | 0 | 0.015 (-0.089 to 0.118) | NA | 0.015 (-0.089 to 0.118) | NA | NA | NA |
| Peramivir vs. Standard care/placebo | 1 | 1 | -0.029 (-0.097 to 0.040) | -0.029 (-0.097 to 0.040) | NA | NA | NA | NA |

Comparison: treatment comparison; k: number of studies providing direct evidence; prop: direct evidence proportion; NMA: estimated treatment effect (RD) in network meta-analysis; direct: estimated treatment effect (RD) derived from direct evidence; indirect: estimated treatment effect (RD) derived from indirect evidence; Diff: difference between direct and indirect treatment estimates; z: z-value of test for disagreement (direct versus indirect): Incoherence p-value: p-value of test for disagreement (direct versus indirect). NA: not applicable.

#### 8.3. Direct, indirect, and network treatment estimates for duration of hospitalization

| **Comparison** | **k** | **Prop** | **NMA (95% CI)** | **Direct (95% CI)** | **Indirect (95% CI)** | **Diff (95% CI)** | **z** | **Incoherence p-value** |
| --- | --- | --- | --- | --- | --- | --- | --- | --- |
| Oseltamivir vs. Peramivir | 1 | 1 | 0.1 (-0.98 to 1.18) | 0.10 (-0.98 to 1.18) | NA | NA | NA | NA |
| Oseltamivir vs. Standard care/placebo | 2 | 1 | -1.63 (-2.81 to -0.45) | -1.63 (-2.81 to -0.45) | NA | NA | NA | NA |
| Peramivir vs. Standard care/placebo | 0 | 0 | -1.73 (-3.33 to -0.13) | NA | -1.73 (-3.33 to -0.13) | NA | NA | NA |

Comparison: treatment comparison; k: number of studies providing direct evidence; prop: direct evidence proportion; NMA: estimated treatment effect (MD) in network meta-analysis; direct: estimated treatment effect (MD) derived from direct evidence; indirect: estimated treatment effect (MD) derived from indirect evidence; Diff: difference between direct and indirect treatment estimates; z: z-value of test for disagreement (direct versus indirect): Incoherence p-value: p-value of test for disagreement (direct versus indirect). NA: not applicable.

#### 8.4. Direct, indirect, and network treatment estimates for time to alleviation of symptoms

| **Comparison** | **k** | **Prop** | **NMA (95% CI)** | **Direct (95% CI)** | **Indirect (95% CI)** | **Diff (95% CI)** | **z** | **Incoherence p-value** |
| --- | --- | --- | --- | --- | --- | --- | --- | --- |
| Oseltamivir vs. Peramivir | 2 | 1 | 0.39 (-0.63 to 1.40) | 0.39 (-0.63 to 1.40) | NA | NA | NA | NA |
| Oseltamivir vs. Standard care/placebo | 0 | 0 | 0.34 (-0.86 to 1.54) | NA | 0.34 (-0.86 to 1.54) | NA | NA | NA |
| Peramivir vs. Standard care/placebo | 1 | 1 | -0.05 (-0.69 to 0.59) | -0.05 (-0.69 to 0.59) | NA | NA | NA | NA |

Comparison: treatment comparison; k: number of studies providing direct evidence; prop: direct evidence proportion; NMA: estimated treatment effect (MD) in network meta-analysis; direct: estimated treatment effect (MD) derived from direct evidence; indirect: estimated treatment effect (MD) derived from indirect evidence; Diff: difference between direct and indirect treatment estimates; z: z-value of test for disagreement (direct versus indirect): Incoherence p-value: p-value of test for disagreement (direct versus indirect). NA: not applicable.

#### 8.5. Direct, indirect, and network treatment estimates for any adverse events

| **Comparison** | **k** | **Prop** | **NMA (95% CI)** | **Direct (95% CI)** | **Indirect (95% CI)** | **RoR (95% CI)** | **z** | **Incoherence p-value** |
| --- | --- | --- | --- | --- | --- | --- | --- | --- |
| Oseltamivir vs. Peramivir | 1 | 1 | 0.77 (0.52 to 1.14) | 0.77 (0.52 to 1.14) | NA | NA | NA | NA |
| Oseltamivir vs. Zanamivir | 1 | 1 | 1.12 (0.99 to 1.28) | 1.12 (0.99 to 1.28) | NA | NA | NA | NA |
| Peramivir vs. Zanamivir | 0 | 0 | 1.46 (0.97 to 2.21) | NA | 1.46 (0.97 to 2.21) | NA | NA | NA |

Comparison: treatment comparison; k: number of studies providing direct evidence; prop: direct evidence proportion; NMA: estimated treatment effect (RR) in network meta-analysis; direct: estimated treatment effect (RR) derived from direct evidence; indirect: estimated treatment effect (RR) derived from indirect evidence; RoR: Ratio of Ratios (direct versus indirect); z: z-value of test for disagreement (direct versus indirect): Incoherence p-value: p-value of test for disagreement (direct versus indirect). NA: not applicable.

#### 8.6. Direct, indirect, and network treatment estimates for serious adverse events

| **Comparison** | **k** | **Prop** | **NMA (95% CI)** | **Direct (95% CI)** | **Indirect (95% CI)** | **RoR (95% CI)** | **z** | **Incoherence p-value** |
| --- | --- | --- | --- | --- | --- | --- | --- | --- |
| Oseltamivir vs. Peramivir | 1 | 1 | 0.79 (0.26 to 2.39) | 0.79 (0.26 to 2.39) | NA | NA | NA | NA |
| Oseltamivir vs. Zanamivir | 1 | 1 | 1.07 (0.75 to 1.53) | 1.07 (0.75 to 1.53) | NA | NA | NA | NA |
| Peramivir vs. Zanamivir | 0 | 0 | 1.35 (0.42 to 4.32) | NA | 1.35 (0.42 to 4.32) | NA | NA | NA |

Comparison: treatment comparison; k: number of studies providing direct evidence; prop: direct evidence proportion; NMA: estimated treatment effect (RR) in network meta-analysis; direct: estimated treatment effect (RR) derived from direct evidence; indirect: estimated treatment effect (RR) derived from indirect evidence; RoR: Ratio of Ratios (direct versus indirect); z: z-value of test for disagreement (direct versus indirect): Incoherence p-value: p-value of test for disagreement (direct versus indirect). NA: not applicable.

### Appendix 9. GRADE summary of findings for outcomes

#### 9.1. GRADE summary of findings for admission to ICU for different comparisons

| **Comparison** | **Study results and measurements** | **Absolute difference (95% CI)** | **Certainty in effect estimates** | **Plain language summary** |
| --- | --- | --- | --- | --- |
| Oseltamivir versus Standard care/placebo | Risk difference: 0.015 (95% CI -0.089 to 0.118) Based on indirect evidence | 15 more per 1000  (95% CI 89 fewer to 118 more) | Very low†* | Whether oseltamivir reduces admission to ICU is very uncertain. |
| Peramivir versus Standard care/placebo | Risk difference: -0.029  (95% CI -0.097 to 0.040) Based on data from 98 participants in 1 study | 29 fewer per 1000  (95% CI 97 fewer to 40 more) | Very low†‡ | Whether peramivir reduces admission to ICU is very uncertain. |
| Oseltamivir versus Peramivir | Risk difference: 0.043 (95% CI -0.034 to 0.121) Based on data from 137 participants in 1 study | 43 more per 1000  (95% CI 34 fewer to 121 more) | Very low* | Whether oseltamivir reduces admission to ICU compared with peramivir is very uncertain. |

*Rated down 3 levels for imprecision.

†Rated down 1 level for risk of bias.

‡Rated down 2 levels for imprecision.

#### 9.2. GRADE summary of findings for time to alleviation of symptoms for different comparisons

| **Comparison** | **Mean difference (95% CI)** | **Certainty in effect estimates** | **Plain language summary** |
| --- | --- | --- | --- |
| Oseltamivir versus Standard care/placebo | 0.34 (-0.86 to 1.54) | Low†‡ | Oseltamivir may have little or no effect on time to alleviation of symptoms. |
| Peramivir versus Standard care/placebo | -0.05 (-0.69 to 0.59) | Low†‡ | Peramivir may have little or no effect on time to alleviation of symptoms. |
| Oseltamivir versus Peramivir | 0.39 (-0.63 to 1.40) | Low†‡ | There may be little or no difference between oseltamivir and peramivir in time to alleviation of symptoms. |

†Rated down 1 level for risk of bias.

‡Rated down 1 level for imprecision.

#### 9.3. GRADE summary of findings for any adverse events for different comparisons

| **Comparison** | **Study results and measurements** | **Absolute effect estimates (per 1000)** | | **Absolute difference (95% CI)** | **Certainty in effect estimates** | **Plain language summary** |
| --- | --- | --- | --- | --- | --- | --- |
| Oseltamivir versus Peramivir | Relative risk: 0.77  (95% CI 0.52 to 1.14)  Based on data from 137 participants in 1 study | Peramivir: 851 | Oseltamivir: 655 | 196 fewer per 1000 (95% CI 408 fewer to 119 more) | Very low†‡ | Whether oseltamivir increases any adverse events compared with peramivir is very uncertain. |
| Oseltamivir versus Zanamivir | Relative risk: 1.12  (95% CI 0.99 to 1.28)  Based on data from 615 participants in 1 study | Zanamivir: 583 | Oseltamivir: 653 | 70 more per 1000 (95% CI 6 fewer to 163 more) | Very low†‡ | Whether oseltamivir increases any adverse events compared with zanamivir is very uncertain. |
| Peramivir versus Zanamivir | Relative risk: 1.46  (95% CI 0.97 to 2.21)  Based on indirect evidence | Zanamivir: 583 | Peramivir: 851 | 268 more per 1000 (95% CI 17 fewer to 417 more) | Very low†‡ | Whether peramivir increases any adverse events compared with zanamivir is very uncertain. |

†Rated down 1 level for risk of bias.

‡Rated down 2 levels for imprecision.

#### 9.4. GRADE summary of findings for serious adverse events for different comparisons

| **Comparison** | **Study results and measurements** | **Absolute effect estimates (per 1000)** | | **Absolute difference (95% CI)** | **Certainty in effect estimates** | **Plain language summary** |
| --- | --- | --- | --- | --- | --- | --- |
| Oseltamivir versus Peramivir | Relative risk: 0.79  (95% CI 0.26 to 2.39)  Based on data from 137 participants in 1 study | Peramivir: 234 | Oseltamivir: 185 | 49 fewer per 1000 (95% CI 173 fewer to 325 more) | Very low†‡ | Whether oseltamivir increases serious adverse events compared with peramivir is very uncertain. |
| Oseltamivir versus Zanamivir | Relative risk: 1.07  (95% CI 0.75 to 1.53)  Based on data from 615 participants in 1 study | Zanamivir: 173 | Oseltamivir: 185 | 12 more per 1000 (95% CI 43 fewer to 92 more) | Very low†‡ | Whether oseltamivir increases serious adverse events compared with zanamivir is very uncertain. |
| Peramivir versus Zanamivir | Relative risk: 1.35  (95% CI 0.42 to 4.32)  Based on indirect evidence | Zanamivir: 173 | Peramivir: 234 | 61 more per 1000 (95% CI 100 fewer to 574 more) | Very low†‡ | Whether peramivir increases serious adverse events compared with zanamivir is very uncertain. |

†Rated down 1 level for risk of bias.

‡Rated down 2 levels for imprecision.

#### 9.5. GRADE summary of findings for progression to mechanical ventilation, emergence of resistance, and adverse events related to treatments serious adverse events

| **Outcomes** | **Comparison** | **Study results and measurements** | **Absolute effect estimates (per 1000)** | | **Absolute difference (95% CI)** | **Certainty in effect estimates** | **Plain language summary** |
| --- | --- | --- | --- | --- | --- | --- | --- |
| Progression to mechanical ventilation | Oseltamivir versus Zanamivir | Relative risk: 1.20  (95% CI 0.90 to 1.62)  Based on data from 488 participants in 1 study | Zanamivir: 255 | Oseltamivir: 306 | 51 more per 1000 (95% CI 26 fewer to 158 more) | Very low†‡ | Whether oseltamivir reduces progression to mechanical ventilation compared with zanamivir is very uncertain. |
| Emergence of resistance | Oseltamivir versus Zanamivir | Relative risk: 2.89  (95% CI 0.88 to 9.49)  Based on data from 615 participants in 1 study | Zanamivir: 10 | Oseltamivir: 29 | 19 more per 1000 (95% CI 1 fewer to 85 more) | Very low†‡ | Whether oseltamivir increases emergence of resistance compared with zanamivir is very uncertain. |
| Adverse events related to treatments | Oseltamivir versus Zanamivir | Relative risk: 1.49  (95% CI 1.00 to 2.23)  Based on data from 615 participants in 1 study | Zanamivir: 115 | Oseltamivir: 171 | 56 more per 1000 (95% CI 0 fewer to 141 more) | Very low†‡ | Whether oseltamivir increases adverse events related to treatments compared with zanamivir is very uncertain. |

†Rated down 1 level for risk of bias.

‡Rated down 2 levels for imprecision.

#### 9.6. GRADE summary of findings for duration of mechanical ventilation

| **Comparison** | **Mean difference (95% CI)** | **Certainty in effect estimates** | **Plain language summary** |
| --- | --- | --- | --- |
| Oseltamivir versus Zanamivir | 0.89 (-2.32 to 4.10) | Very low†‡ | Whether oseltamivir reduces duration of mechanical ventilation compared with zanamivir is very uncertain. |

†Rated down 1 level for risk of bias.

‡Rated down 2 levels for imprecision.

### Appendix 10. Results of a study, comparing baloxavir plus NAIs with NAIs, not included in the network meta-analysis

#### 10.1. Forest plots for baloxavir plus NAIs versus NAIs (oseltamivir, zanamivir, or peramivir)

Dichotomous outcomes

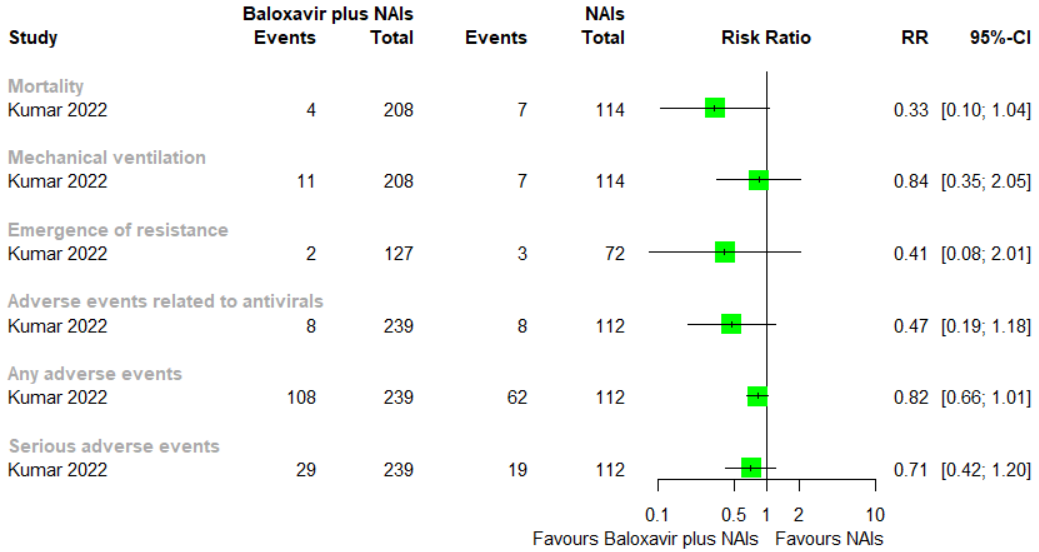

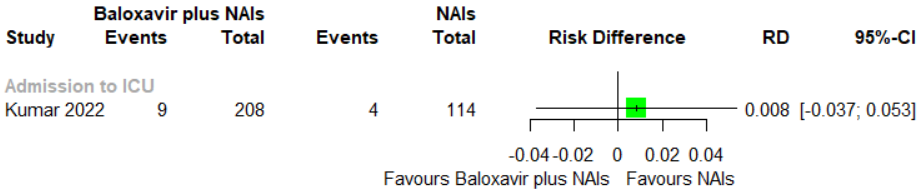

Continuous outcomes

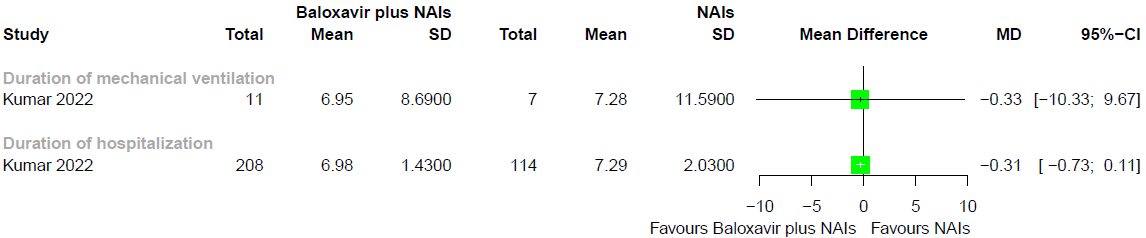

#### 10.2. GRADE summary of findings for baloxavir plus NAIs versus NAIs (oseltamivir, zanamivir, or peramivir)

| **Outcome**  Timeframe | **Study results and measurements** | **Absolute effect estimates** | | **Certainty of the Evidence**  (Quality of evidence) | **Summary** |
| --- | --- | --- | --- | --- | --- |
|  |  | NAIs | Baloxavir plus NAIs |  |  |
| Mortality (Seasonal influenza) | Relative risk: 0.33  (95% CI 0.10 - 1.04)  Based on data from 322 participants in 1 study | **15**  per 1000 | **5**  per 1000 | **Very low**  Due to extremely serious imprecision^1^ | Whether baloxavir plus NAIs reduces mortality in people with seasonal influenza compared with NAIs is very uncertain. |
|  |  | Difference: **10 fewer per 1000**  (95% CI 13 fewer - 1 more) | |  |  |
| Mortality (Zoonotic influenza) | Relative risk: 0.33  (95% CI 0.1 - 1.04)  Based on data from 322 participants in 1 study | **195**  per 1000 | **64**  per 1000 | **Very low**  Due to extremely serious imprecision^1^ | Whether baloxavir plus NAIs reduces mortality in people with zoonotic influenza compared with NAIs is very uncertain. |
|  |  | Difference: **131 fewer per 1000**  (95% CI 175 fewer - 8 more) | |  |  |
| Admission to ICU | Risk difference: 0.008  (95% CI -0.037 - 0.053)  Based on data from 322 participants in 1 study | **35**  per 1000 | **43**  per 1000 | **Very low**  Due to extremely serious imprecision^1^ | Whether baloxavir plus NAIs reduces admission to ICU compared with NAIs is very uncertain. |
|  |  | Difference: **8 more per 1000**  (95% CI 37 fewer - 53 more) | |  |  |
| Mechanical ventilation | Relative risk: 0.84  (95% CI 0.35 - 2.05)  Based on data from 322 participants in 1 study | **61**  per 1000 | **51**  per 1000 | **Very low**  Due to extremely serious imprecision^1^ | Whether baloxavir plus NAIs reduces mechanical ventilation compared with NAIs is very uncertain. |
|  |  | Difference: **10 fewer per 1000**  (95% CI 40 fewer - 64 more) | |  |  |
| Any adverse events | Relative risk: 0.82  (95% CI 0.66 - 1.01)  Based on data from 351 participants in 1 study | **554**  per 1000 | **454**  per 1000 | **Very low**  Due to extremely serious imprecision^1^ | Whether baloxavir plus NAIs increases any adverse events compared with NAIs is very uncertain. |
|  |  | Difference: **100 fewer per 1000**  (95% CI 188 fewer - 6 more) | |  |  |
| Adverse events related to treatment | Relative risk: 0.47  (95% CI 0.19 - 1.18)  Based on data from 351 participants in 1 study | **71**  per 1000 | **33**  per 1000 | **Very low**  Due to extremely serious imprecision^1^ | Whether baloxavir plus NAIs increases adverse events related to treatment compared with NAIs is very uncertain. |
|  |  | Difference: **38 fewer per 1000**  (95% CI 58 fewer - 13 more) | |  |  |
| Serious adverse events | Relative risk: 0.71  (95% CI 0.42 - 1.2)  Based on data from 351 participants in 1 study | **170**  per 1000 | **121**  per 1000 | **Very low**  Due to extremely serious imprecision^1^ | Whether baloxavir plus NAIs increases serious adverse events compared with NAIs is very uncertain. |
|  |  | Difference: **49 fewer per 1000**  (95% CI 99 fewer - 34 more) | |  |  |
| Emergence of resistance | Relative risk: 0.41  (95% CI 0.08 - 2.01)  Based on data from 199 participants in 1 study | **42**  per 1000 | **17**  per 1000 | **Low**  Due to very serious imprecision^2^ | Baloxavir plus NAIs may have little or no effect on emergence of resistance compared with NAIs. |
|  |  | Difference: **25 fewer per 1000**  (95% CI 39 fewer - 42 more) | |  |  |
| Duration of hospitalization | Measured by: day  Lower better  Based on data from 322 participants in 1 study | **7.29**  Mean | **6.98**  Mean | **Low**  Due to very serious imprecision^2^ | Baloxavir plus NAIs may have little or no effect on duration of hospitalization compared with NAIs. |
|  |  | Difference: **MD 0.31 lower**  (95% CI 0.73 lower - 0.11 higher) | |  |  |
| Duration of mechanical ventilation | Measured by: day  Lower better  Based on data from 18 participants in 1 study | **7.28**  Mean | **6.95**  Mean | **Very low**  Due to extremely serious imprecision^1^ | Whether baloxavir plus NAIs reduces duration of mechanical ventilation compared with NAIs is very uncertain. |
|  |  | Difference: **MD 0.33 lower**  (95% CI 10.33 lower - 9.67 higher) | |  |  |

1. **Imprecision: extremely serious.** Wide confidence intervals, only data from one study
2. **Imprecision: very serious.** Only data from one study

### Appendix 11. Results of a study, comparing zanamivir plus rimantadine with rimantadine, not included in the network meta-analysis

#### 11.1. Forest plots for zanamivir plus rimantadine versus rimantadine

Dichotomous outcomes

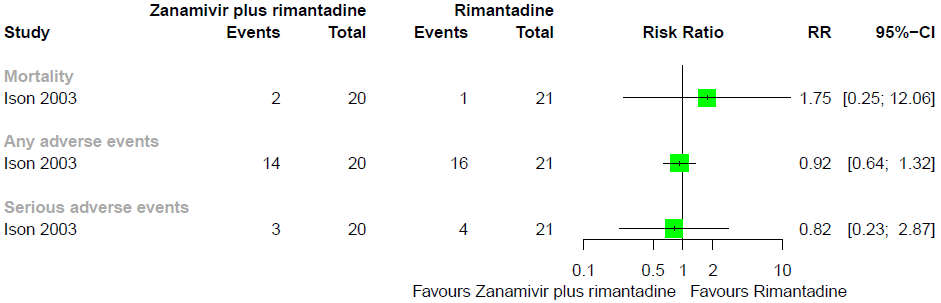

Continuous outcomes- Duration of hospitalization

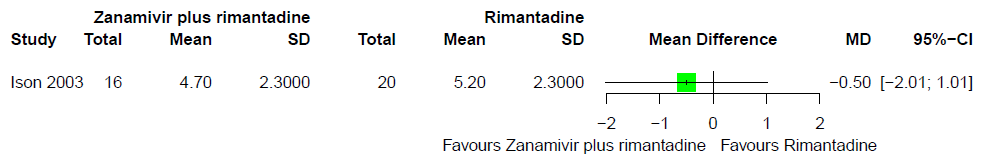

#### 11.2. GRADE summary of findings for zanamivir plus rimantadine versus rimantadine

| **Outcome**  Timeframe | **Study results and measurements** | **Absolute effect estimates** | | **Certainty of the Evidence**  (Quality of evidence) | **Summary** |
| --- | --- | --- | --- | --- | --- |
|  |  | Rimantadine | Zanamivir plus rimantadine |  |  |
| Mortality (Seasonal influenza) | Relative risk: 1.75  (95% CI 0.25 - 12.06)  Based on data from 41 participants in 1 study | **24**  per 1000 | **42**  per 1000 | **Very low**  Due to serious risk of bias, Due to extremely serious imprecision^1^ | Whether zanamivir plus rimantadine reduces mortality in people with seasonal influenza compared with rimantadine is very uncertain. |
|  |  | Difference: **18 more per 1000**  (95% CI 18 fewer - 265 more) | |  |  |
| Mortality (Seasonal influenza) | Relative risk: 1.75  (95% CI 0.25 - 12.06)  Based on data from 41 participants in 1 study | **310**  per 1000 | **543**  per 1000 | **Very low**  Due to serious risk of bias, Due to extremely serious imprecision^1^ | Whether zanamivir plus rimantadine reduces mortality in people with zoonotic influenza compared with rimantadine is very uncertain. |
|  |  | Difference: **233 more per 1000**  (95% CI 232 fewer - 690 more) | |  |  |
| Any adverse events | Relative risk: 0.92  (95% CI 0.64 - 1.32)  Based on data from 41 participants in 1 study | **762**  per 1000 | **701**  per 1000 | **Very low**  Due to serious risk of bias, Due to extremely serious imprecision^1^ | Whether zanamivir plus rimantadine increases any adverse events compared with rimantadine is very uncertain. |
|  |  | Difference: **61 fewer per 1000**  (95% CI 274 fewer - 244 more) | |  |  |
| Serious adverse events | Relative risk: 0.82  (95% CI 0.23 - 2.87)  Based on data from 41 participants in 1 study | **190**  per 1000 | **156**  per 1000 | **Very low**  Due to serious risk of bias, Due to extremely serious imprecision^1^ | Whether zanamivir plus rimantadine increases any adverse events compared with rimantadine is very uncertain. |
|  |  | Difference: **34 fewer per 1000**  (95% CI 146 fewer - 355 more) | |  |  |
| Duration of hospitalization | Measured by: day  Lower better  Based on data from 36 participants in 1 study | **5.20**  Mean | **4.70**  Mean | **Very low**  Due to serious risk of bias, Due to very serious imprecision^2^ | Whether zanamivir plus rimantadine reduces duration of hospitalization compared with rimantadine is very uncertain. |
|  |  | Difference: **MD 0.50 lower**  (95% CI 2.01 lower - 1.01 higher) | |  |  |

1. **Risk of Bias: serious.** Trials stopping earlier than scheduled, resulting in potential for overestimating benefits, Incomplete data and/or large loss to follow up; **Imprecision: extremely serious.** Very wide confidence intervals, only data from one study
2. **Risk of Bias: serious.** Trials stopping earlier than scheduled, resulting in potential for overestimating benefits, Incomplete data and/or large loss to follow up; **Imprecision: very serious.** Wide confidence intervals, only data from one study
